# A multi-omics approach identifies a blood-based miRNA signature of cognitive decline in two large observational trials

**DOI:** 10.1101/2022.06.17.22276532

**Authors:** Angélique Sadlon, Petros Takousis, Evangelos Evangelou, Inga Prokopenko, Panagiotis Alexopoulos, Chinedu T Udeh-Momoh, Geraint Price, Lefkos Middleton, Robert Perneczky, the Alzheimer’s Disease Neuroimaging Initiative

## Abstract

Identifying individuals before the onset of overt symptoms is a key prerequisite for the prevention of Alzheimer’s disease (AD). A wealth of data reports dysregulated microRNA (miRNA) expression in the blood of individuals with AD, but evidence in individuals at subclinical stages is sparse. In this study, a qPCR analysis of a prioritised set of 38 candidate miRNAs in the blood of 830 healthy individuals from the CHARIOT PRO cohort (West London, UK) was undertaken. Here, we identified six differentially expressed miRNAs (hsa-miR-128-3p, hsa-miR-144-5p, hsa-miR-146a-5p, hsa-miR-26a-5p, hsa-miR-29c-3p and hsa-miR-363-3p) in the blood of individuals with low cognitive performance on the Repeatable Battery for the Assessment of Neuropsychological Status (RBANS). A pathway enrichment analysis for the six miRNAs indicated involvement of apoptosis and inflammation, relevant in early AD stages. Subsequently, we used whole genome sequencing (WGS) data from 750 individuals from the AD Neuroimaging Initiative (ADNI) to perform a genetic association analysis for polymorphisms within the significant miRNAs’ genes and CSF concentrations of phosphorylated-tau, total-tau, amyloid-β42 and soluble-TREM2 and BACE1 activity. Our analysis revealed 24 SNPs within *MIR29C* to be associated with CSF levels of amyloid-β42 and soluble-TREM2 and BACE1 activity. Our study shows the potential of a six-miRNA set as diagnostic blood biomarker of subclinical cognitive deficits in AD. Polymorphisms within *MIR29C* suggest a possible interplay between the amyloid cascade and microglial activation at preclinical stages of AD.

## Introduction

Alzheimer’s disease (AD) is the most frequent neurodegenerative disorder, accounting for two thirds of dementia cases. Forecasting models project a dramatic increase of dementia prevalence in the coming years, with expected 152 million affected individuals worldwide by 2050 ^1^. AD drug development has experienced a high failure rate and a disease modifying therapy with unequivocally proven effectiveness for AD is yet to be developed ^2^. Currently approved drugs for AD are aimed at symptomatic individuals, i.e., those in the mild cognitive impairment (MCI) or dementia stage, in whom neuronal loss may be too advanced to be slowed down or stopped. Therefore, preclinical AD is receiving increased attention as a window of opportunity for therapies aiming at slowing disease progression ^3^.

AD pathology may start several decades before the onset of overt symptoms, accompanied by characteristic biomarker changes ^4, 5^, including increased cerebrospinal (CSF) levels of phosphorylated-tau (p-tau)217, p-tau181 and total-tau (t-tau), in both familial and sporadic AD ^6, 7^, and increased amyloid-β (Aβ) and tau radiotracer uptake on positron-emission-tomography (PET) ^8, 9^. Blood levels of p-tau181 (and other phosphorylation states) show promise as biomarker candidates for predicting AD dementia in cognitively unimpaired individuals ^10, 11^, but they remain to be validated fully. All these biomarkers rely either on invasive methods, exposure to radiation, or techniques not available widely, limiting their usefulness in routine population settings.

Blood microRNAs (miRNAs) are attracting increased interest as novel minimally invasive biomarker candidates for AD. First discovered in the early 2000s, miRNAs are small noncoding molecules, regulating gene expression at a post-transcriptional level ^12^. A large number of miRNAs participate in several pathways altered in AD, such as apoptosis, inflammatory signalling or protein degradation ^13^. MiRNAs can be measured in any body fluid or tissue, and we have recently reported a consistent and reproducible dysregulated expression in the CSF, brain and blood of patients with AD ^14^. Moreover, measurement of selected miRNAs in the blood shows good performance in differentiating healthy controls from individuals with AD ^15^. Recently, a signature of three miRNAs (miR-181a, miR-148a and miR-146a) was reported to correlate with cognitive function in cognitively healthy individuals as well as with cognitive decline in a mouse model. The authors also reported increased blood levels of these three miRNAs in individuals progressing from MCI to AD within two years ^16^.

Interestingly, few studies have investigated the role of miRNAs in preclinical stages of AD. Yet, several miRNAs are directly involved in pathways initiating early pathological changes. For instance, miR-146a, downregulated in the blood of AD patients, represses the expression of neurofilament light chains (NfL), increased several years before the onset of clinical symptoms in individuals carrying a mutation in the *APP, PSEN1 or PSEN2* familial AD genes ^17, 18^. Similarly, miR-125b, also downregulated in AD, blocks the translation of SphK1, a mediator of neuroinflammation in early stages ^19, 20^. In parallel, the development of large-scale genome wide analysis techniques has offered novel insights into the role of miRNA gene polymorphisms in the development of neurological disorders such as Tourette syndrome ^21^. Mutations within a miRNA gene may affect different stages of its transcript processing and may result in abrogated miRNA function^22^. In AD, the presence of rs2910164 within *MIR146A* resulted in decreased levels of miR-146a-5p, correlating with increased levels of TLR-2, a critical microglial receptor mediating neuroinflammation following its binding to Aβ42 ^23, 24^. Similarly, the presence of rs6070628 within *MIR298* correlated with higher CSF levels of p-tau181 in a dose-dependent fashion ^25^.

The aim of this study was to investigate the expression of selected blood miRNAs in healthy subjects at different level of cognitive performance. We prioritised significantly dysregulated miRNAs in AD based on our recent systematic review and meta-analysis of the literature ^14^. In subsequent steps to provide a biological validation of our miRNA targets, we performed pathway enrichment analyses and explored associations between SNPs within genes coding for miRNAs dysregulated in AD and common CSF markers of neurodegeneration were investigated.

## Material and methods

### Study populations

For the miRNA expression analyses, we accessed the biobanked samples of the prospective, observational CHARIOT PRO Main Study cohort (Cognitive Health in Ageing Register: Investigational, Observational, and Trial studies in dementia research: Prospective Readiness cOhort, clinicaltrials.gov ID NCT02114372), recruited in West London (UK) between February 2014 and December 2016. The study was terminated in 2017 and was replaced by the still on-going prospective longitudinal biomarker-enriched CHARIOT PRO Sub-Study ^26^. Subjects were aged 60 to 85 and were at different levels of cognitive performance based on their Repeatable Battery for the Assessment of Neuropsychological Status (RBANS) score at baseline. Exclusion criteria included a dementia or MCI diagnosis, presence of any neurological or psychiatric condition, substance use disorder or the presence of reversible causes of dementia. Further details can be found elsewhere ^27^.

For the genetic association analysis with CSF biomarkers, we downloaded data from the AD Neuroimaging Initiative (ADNI) database. ADNI was first launched in 2003 as part of a public-private partnership to investigate the role of imaging and biological biomarkers in the progression of MCI to AD (www.adni-info.org). The ADNI cohort includes subjects with AD dementia, MCI and healthy controls ^28^. AD dementia was diagnosed based on the National Institute of Neurological and Communicative Disorders and Stroke-AD and Related Disorders Association (NINCDS-ADRDA) criteria ^29^. Individuals with AD had a Mini-Mental State Examination (MMSE) score of 20-26 and a Clinical Dementia Rating (CDR) score of 0.5 or 1. Individuals without functional complaints but a MMSE score of 24-30, a CDR score of 0.5 (with a memory box score of 0.5 or greater) and memory complaints were classified as MCI. Healthy controls were free of memory complaints, impairment on cognitive testing and were independent in their activities of daily living ^30^. For both cohorts, subjects provided written informed consent.

### Ethics approval

CHARIOT PRO was conducted in accordance with Good Clinical Practice (GCP) Guidelines, Guidelines for Good Pharmacovigilance Practices (GPP) issued by the International Society for Pharmacoepidemiology (ISPE), applicable national guidelines, and to the Declaration of Helsinki. An independent ethics committee approved participant written informed consent forms before enrolment collected during the baseline clinic visit.

ADNI was reviewed and approved by all host study site institutional review boards and participants completed informed consent after receiving a comprehensive description of the ADNI. All procedures performed in studies involving human participants were in accordance with the ethical standards of the institutional and/or national research committee and with the 1964 Helsinki declaration and its later amendments or comparable ethical standards. Informed consent: Informed consent was obtained from all individual participants included in the study.

### Neuropsychological measurements in CHARIOT PRO

The RBANS score was used to measure cognitive function in the CHARIOT PRO cohort. Briefly, this method assesses five cognitive domains (immediate memory, visuospatial/ constructional, language, attention and delayed memory) using 12 tests ^31^. The individual five tested domains are summed in a total score, subsequently adjusted to a normative dataset, including 540 healthy subjects 20-89 years old ^32^. The RBANS was validated in community dwelling individuals and is used as a screening tool for dementia in clinical practice and trials ^33^ In a previous study, 1.5 standard deviation (SD) below the mean performed best when differentiating MCI patients from healthy controls, while 2 SD below the mean identified individuals with more advanced stages and functional impairment. Furthermore, the lower end of normal performance was identified as 1 SD below the mean ^34^. Therefore, we defined subtle cognitive deficits in overall cognitively normal individuals using a cut-off of 1 SD below the mean total RBANS score.

### qPCR analysis in CHARIOT PRO

In total, 46 miRNAs were analysed, including 38 prioritised candidate miRNAs (32 in the blood and six in the brain) ^14^ selected based on our recent systematic review and meta-analysis of over 100 studies and another eight endogenous miRNAs with reportedly stable expression in the blood ^35^. Two spike-in controls, UniSp3 and UniSp6, were used as serial quality controls throughout the qPCR process. Blood samples were collected in PAXgene Blood RNA tubes (Qiagen, Venlo, The Netherlands). Subsequent qPCR analyses were performed on the miRCURY LNA miRNA PCR System at Qiagen laboratories. Further details can be found in the supplementary material.

During quality control of the qPCR data, qPCR cycle threshold (Ct) values > 35 (i.e., the number of amplification cycles needed for the target to be detected above the background signal) were considered not available, according to the manufacturer’s recommendations. MiRNAs with Ct values > 35 in more than 50% of the samples were removed (total miRNAs removed: n=18 (39.13%), which is comparable to other studies ^36^) (Supplementary material table 1). Finally, normalization was undertaken using the *geNorm* algorithm, preferred over other normalization approaches according to a recent comparative study ^36^. In a final step, Ct values were converted to fold changes.

### MiRNA gene region definition

The region of interest was defined 200 kb before and after the start of the miRNA gene based on the GRCh37 genome assembly. Haplotype blocks were identified using Haploview and applying the Gabriel et al algorithm ^37, 38^. This method uses D prime, a marker of linkage disequilibrium (LD), between two SNPs to identify a haplotype block defined as the presence of 95% SNPs in strong LD. A pair of SNPs are in strong LD having the one-sided upper 95% confidence bound on D’ is >0.98 (consistent with no historical recombination) and the lower bound D’>0.7.

### Whole genome sequencing in ADNI

Whole genome sequencing (WGS) data were provided by the ADNI genetics core team. The analyses were undertaken using the Illumina HiSeq2000 platform and followed the Genome Analysis Toolkit (GATK) pipeline. Full description of the methods is available elsewhere ^39^. Following download of the ADNI data in PLINK format, we conducted additional quality control procedures based on a previously described protocol ^40^. In brief, after a check of discordant sex information, we removed individuals with high missing and outlying heterozygosity rates (cut-off call rate > 0.03, cut-off heterozygosity > 3 SD), duplicated and related individuals (cut-off Pi-hat 0.2). To avoid potential biases from population stratification we performed a principal component analysis (PCA) using data from the 1,000 Genomes Project and individuals with European ancestry were retained.

### Biomarker measurements in ADNI

CSF biomarker measurements and associated data were downloaded from ADNI. All analyses followed ADNI standard operating procedures and published protocols. Briefly, Aβ42, t-tau and p-tau181 analyses were performed on the multiplex xMAP platform (Luminex Corp., Austin, TX, USA) with monoclonal antibodies specific for Aβ42 (4D7A3), t-tau (AT120), and p-tau181p (AT270) ^41^. A sandwich enzyme-linked immunosorbent assay (ELISA) was used to measure NfL levels in the CSF with a cut off for quantification of 50 ng/L ^42^. Soluble triggering receptor expressed on myeloid cells 2 (sTREM2) was measured using a previously described ELISA protocol ^43, 44^. Finally, beta-site amyloid precursor protein cleaving enzyme 1 (BACE1) activity levels in the CSF were measured using a two-step assay described previously ^45^.

### Statistical analysis

Distribution of continuous variables was assessed for normality using the Kolmogorov Smirnov test and log10 transformed when appropriate. Demographic differences between the RBANS low performance and normal performance groups were evaluated with a Student’s t-test. Mann-Whitney test was used for non-normally distributed variables and Pearson’s Chi Square test for categorical variables.

For the qPCR analysis, we used partial correlation (Pearson correlation for normally distributed values and Spearman rank correlation for non-normally distributed values) adjusted for age and sex to investigate the relationship between miRNA concentration and RBANS score. Subsequently, a multivariate regression adjusted for age, sex, education status (<13 education years, ≥13 education years), ethnicity and apolipoprotein E (APOE) ε4 carrier status (defined as the presence of at least one ε4 allele) was applied. Using the same covariates, we assessed differences in miRNA concentrations between the two RBANS performance groups using ANCOVA. Finally, for the miRNAs showing a significant difference between the two performance groups we constructed a ROC (receiver operating characteristic) curve and calculated their AUC (area under the curve) to investigate the discrimination ability of the miRNAs between the groups. Confidence intervals of 95% were considered. These analyses were undertaken in R v4.1.2. Baseline characteristics are described as mean ± SD.

Genetic association analysis for CSF biomarkers was undertaken in PLINK v1.07 using linear regression considering an additive genetic model. The following covariates were added in the model: SNP, age, sex, APOEε4-carrier status and diagnosis at baseline. We controlled for population stratification using the first five principal components. To adjust for multiple comparisons, we preferred the Benjamini Hochberg FDR method over the Bonferroni approach as several analysed SNPs were in LD and not independent from each other. Statistical significance was set at FDR α<0.05. Finally, we conducted a Pearson correlation and a multivariate regression analysis (including the above-mentioned covariates) between biomarkers significantly associated with polymorphisms of the same miRNA gene.

### Post-hoc analyses

For the significant miRNAs, we conducted a pathway enrichment analysis following a previously described protocol ^46^. First, we extracted experimentally validated miRNA gene pairs from miRTarbase and to be more AD-specific, selected only genes expressed in the brain according to the Human Protein Atlas (HPA) ^47, 48^. Then, we conducted a functional enrichment in Gene Ontology, KEGG, REACTOME, considering only gene sets with a minimum of 5 and maximum of 500 genes. We then visualised the results using the plugin Enrichment Map in Cytoscape to explore an overlap in significantly enriched gene sets between different miRNAs. The Jaccard and overlap combined coefficient thresholds were set to 0.225. Finally, pathways were grouped manually into families of biological function. Overlap between miRNAs gene targets were visualised using the UpSetR package for R ^49^. For SNPs significantly associated with CSF biomarkers of AD, we undertook a series of post-hoc functional analyses. First, LD analysis was undertaken using the LDmatrix tool, with a default threshold of 0.8 and by selecting only populations of European ancestry ^50^. Second co-localisation of SNPs with expression quantitative trait loci (eQTL) signals were identified by downloading eQTL signals from fresh frozen hippocampal tissue (from the Miami Brain Endowment Bank ™) from the GTex portal ^51^. Third, we used HaploReg v4.1 to obtain regulatory information from the Roadmap Epigenomics project for histone modification marks, chromatin states and DNase hypersensitivity ^52^. The effects on regulatory motifs (also called TF binding sites) were obtained from Chromatin Immunoprecipitation (ChIP) sequencing data from the ENCODE project ^53^. For the functional annotation of SNPs, only information from hippocampus cell lines were selected. Finally, risk SNPs for disease were obtained from VARadb v1.0 which uses a data mining approach over NHGRI GWAS catalogue, GWASdb v2.0, GAD, and GRASP ^54^.

## Results

### Characteristics of the CHARIOT PRO and ADNI cohorts

The CHARIOT PRO cohort included 830 individuals with mean age of 68.70 ± 3.51 years and including 475 (57.23%) female participants (Table 1). The 1SD below the mean cut-off for subtle cognitive deficits (1 SD below the mean) corresponded to an RBANS Total Scale score of 88, and 132 participants (15.90%) fell at or below this level. The two performance groups showed a statistically significant difference in sex (P=0.007), and education status (P=0.008). The ADNI cohort for whom WGS data was available consisted of 812 individuals. After quality control, 62 individuals were removed. Furthermore, among 1 553 534 markers, we excluded a total of 365 876 markers with call rates < 0.9 (96 142 markers) or minor allele frequency (MAF) < 0.05 (269 734 markers). No markers deviated from the Hardy Weinberg Equilibrium at a P < 10^−6^. The sample’s mean age was 73.42 ± 7.03 years, and 322 (42.93%) participants were female (Table 1). For this ADNI sub-cohort, 580, 573, 580, 187, 114, and 123 individuals had data available for Aβ42, t-tau, ptau181, sTREM2, BACE1 and NfL CSF measurements, respectively (Table 1).

**Table 1:**
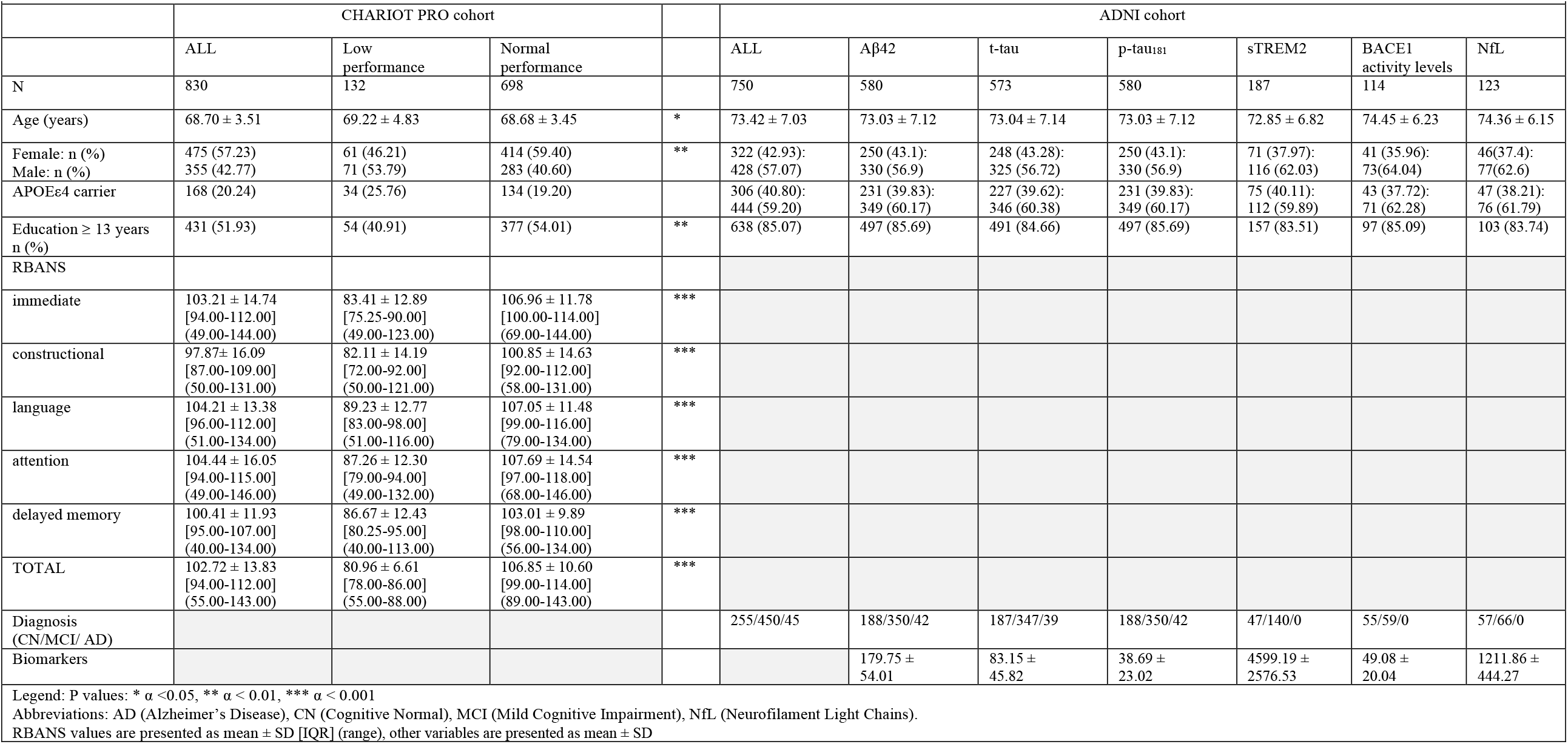
Demographics CHARIOT PRO and ADNI cohorts.

**Table 2:** Significant SNPs overview.

| Table 2: Significant SNPs overview |  |  |  |  |  |  |  |  |
| --- | --- | --- | --- | --- | --- | --- | --- | --- |
|  | SNP name | Position (GRCh37.p13) | strand | allele | Closest annotated gene [distance in kb] | Functional consequence | Regulatory motifs | Diseases |
| **Aβ** | rs7541230 | chr1: 207957555 | + | T | CD46 [0] | intron | Foxj2_2; PLZF; Sox_18; Sox_19 | refractive error, glioma |
| **BACE1** | rs1009897 | chr1: 208063505 | + | G | CD34 [0] | intron, untrsl 5' | NA | hypertension (early onset hypertension), primary rhegmatogenous retinal detachment, glioma |
|  | rs1204682 | chr1: 208077006 | + | G | CD34 [0] | intron | Bbx | primary rhegmatogenous retinal detachment, transmission distortion, infant head circumference |
|  | rs12409962 | chr1: 208044601 | + | T | CD34 [15280] | unknown | YY1_disc2 | Glioma |
|  | rs1555138 | chr1: 208086100 | + | C | CD34 [1416] | near-gene-5 | HDAC2_disc3; NRSF_known1; NRSF_known2; NRSF_known3; Sin3Ak-20_disc7; Znf143_known1 | mitral annular calcium, Parkinson's disease |
|  | rs2250321 | chr1: 208065610 | - | G | CD34 [0] | intron, near-gene-5 | AP-1_known1; BRCA1_known2; Mxi1_disc1; SREBP_disc1; TCF12_disc2 | gene expression of CD34 in dendritic cells, gene expression of CD34 in dendritic cells treated with Mycobacterium tuberculosis |
|  | rs2724373 | chr1: 207999200 | + | C | LOC148696 [3258] | unknown | NA | refractive error, obesity with early age of onset (age >2), general cognitive ability |
|  | rs2785627 | chr1: 208154852 | - | T | PLXNA2 [40734] | unknown | NA | None |
|  | rs2796247 | chr1: 208036976 | + | C | CD34 [22905] | unknown | RXRA_known4 | primary rhegmatogenous retinal detachment, obesity with early age of onset (age >2), glioma |
|  | rs3738460 | chr1: 208071105 | - | A | CD34 [0] | intron | GATA_known1; TAL1_disc1 | Parkinson's disease, years of education |
|  | rs656801 | chr1: 208077643 | + | C | CD34 [0] | intron | Ets_known1 | mitral annular calcium, Parkinson's disease |
|  | rs761277 | chr1: 207993011 | + | A | LOC148696 [0] | intron, ncRNA | Pax-4_4 | gene expression of CD34 in dendritic cells, gene expression of CD34 in dendritic cells treated with Mycobacterium tuberculosis, automobile speeding propensity, glioma |
|  | rs926628 | chr1: 208040165 | + | T | CD34 [19716] | unknown | NA | None |
|  | rs984984 | chr1: 208035088 | + | G | CD34 [24793] | unknown | NA | obesity with early age of onset (age >2), glioma |
| **Aβ and BACE1** | rs2724384 | chr1: 207930203 | + | G | CD46 [0] | Intron | Hoxa5_1; Zfp691 | gene expression of MIR29C in normal prepouch ileum, febrile seizures, immune response to measles vaccine, glioma |
|  | rs2761437 | chr1: 207923081 | + | A | CD46 [2300] | unknown | Hbp1; Mef2_known3 | paternal transmission distortion, glioma |
|  | rs4844390 | chr1: 207934849 | + | A | CD46 [0] | Intron | CTCF_disc1; TCF12_disc2 | glioma |
|  | rs4844623 | chr1: 208035434 | + | T | CD34 [24447] | unknown | Myf_1; PEBP; SEF-1 | glioma |
|  | rs7532674 | chr1: 208026739 | + | G | LOC148696 [30797] | unknown | CCNT2_disc2; NRSF_known3; SP1_disc3; SP2_disc3 | none |
|  | rs882198 | chr1: 208036509 | - | C | CD34 [23372] | unknown | Myf_1; SMC3_disc2 | glioma |
|  | rs7550821 | chr1: 208029947 | + | C | CD34 [29934] | unknown | Foxa_known4; Foxo_2; HNF1_5; Nanog_disc2; Pou5f1_disc1; Pou5f1_known2; Sox_13; Sox_4 | glioma |
| **sTREM2** | rs17041871 | chr1: 208156967 | + | C | PLXNA2 [38619] | unknown | Sox_10; Sox_14; Sox_16; Sox_18; Sox_19; Sox_2; YY1_known6 | None |
|  | rs606714 | chr1: 208151575 | + | G | PLXNA2 [44011] | unknown | CEBPA_2; Foxa_known4; Foxf1; Foxf2; Foxl1_1; Foxo_1; Foxo_2; Foxq1; Hsf_disc1; TEF | None |
|  | rs671273 | chr1: 208145061 | + | A | PLXNA2 [50525] | unknown | CTCF_known1; Pax-4; Rad21_disc1 | None |

Legend: intron variant occurring within an intron, near-gene-5: variant located 5' of a gene, untsl-5: a UTR variant of the 5' UTR

### miRNA and cognitive performance in CHARIOT PRO

In the unadjusted analysis, 17 miRNAs were correlated with the RBANS index scores or the total scale. In a multivariate regression analysis, adjusted for age, sex, education years, ethnicity and APOEε4 carrier status, the RBANS language index was significantly (FDR α = 0.05) associated with hsa-let-7a-5p, hsa-let-7c-5p, hsa-let-7d-5p, hsa-miR-144-5p, hsa-miR-93-5p and hsa-miR-98-5p. Furthermore, we found hsa-miR-363-3p to be significantly (FDR α = 0.05) associated with the RBANS attention index. Finally, hsa-miR-144-5p was significantly (FDR α = 0.05) associated with the RBANS total scale (Supplementary material table 2). When comparing miRNA expression levels between the two RBANS performance groups, six miRNAs (hsa-miR-128-3p, hsa-miR-144-5p, hsa-miR-146a-5p, hsa-miR-26a-5p, hsa-miR-29c-3p, hsa-miR-363-3p) were downregulated in the low performance group compared to the normal performance group (Figure 1A). A significant direct correlation (ranging from rs = 0.09 to rs = 0.55) was found between the six miRNAs (Figure 1B and supplementary material table 3). It is noteworthy that the six miRNAs had similar diagnostic performance, with the hsa-miR-363-3p ROC curve having the highest AUC [95% CI] = 0.761 [0.711-0.812] (Figure 1C).

**Figure 1.**
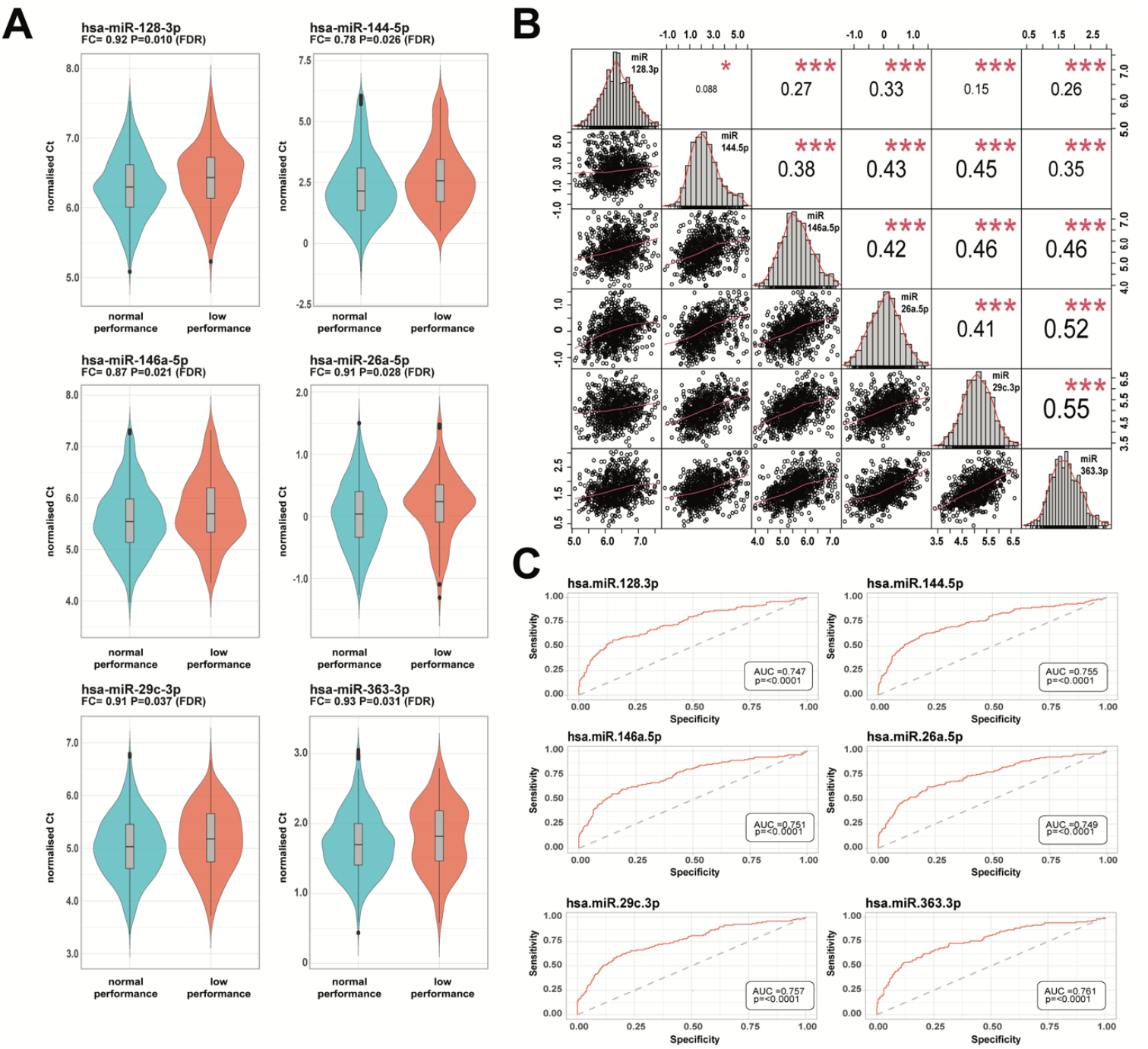
MiRNA expression levels in healthy individuals in CHARIOT PRO (n=830). A) miRNAs significantly dysregulated between the normal vs low performance group B) correlation matrix for the significant miRNAs C) ROC curves for significant miRNAs. Legend: In figure C, the ROC p value reflects the significance level of the deviation from the null hypothesis (i.e. AUC = 0.50)

### Association analysis with CSF biomarkers in ADNI

All six miRNA genes were located on different chromosomes and had a median length of 84.00 (9.25) kb and featured no SNPs with a MAF > 0.05 within them. We identified 40 genes located within 200 kb of these six miRNA genes (Supplementary material table 4). We found significant association for SNPs located within *MIR29C*. Eight SNPs were significantly associated with CSF levels of Aβ42, and minor allele carriers showed lower levels of Aβ42 in the CSF. Moreover, we identified 20 SNPs to be significantly associated with BACE1 activity levels in the CSF. Notably, seven of these SNPs were also significantly associated with CSF levels of Aβ42 and minor allele carriers reported both decreased CSF levels of Aβ42 and BACE1 activity. Finally, we found three SNPs to be significantly associated with sTREM2 levels in the CSF with minor allele carriers showing decreased sTREM2 activity levels (Figure 2A). The SNPs within *MIR29C* associated with Aβ42, BACE1 activity and sTREM2 levels in the CSF were grouped into four haplotype blocks (Supplementary material figure 1). In order to further explore the relationship between CSF levels of Aβ42, BACE1 activity and sTREM2, we undertook a multivariate regression analysis and found a positive correlation between log BACE1 activity (r=0.41, p<0.0001) and log sTREM2 levels in the CSF (β=0.34, 95% CI [0.18,0.49]): for a 10% increase in BACE1 activity in the CSF; the sTREM2 levels in the CSF increased by 3.3%.

**Figure 2.**
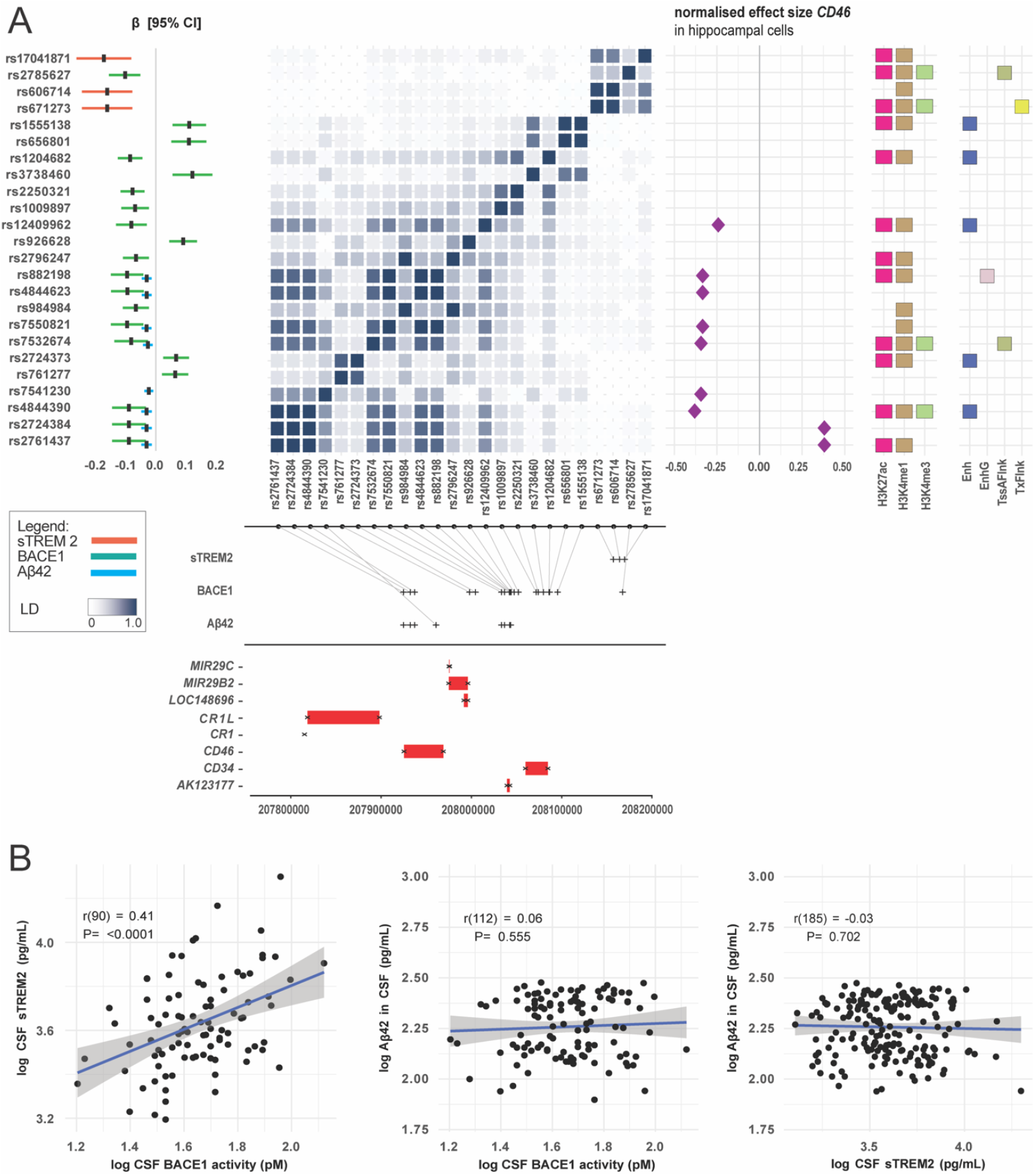
Genetic association results for significant miRNA genes in ADNI. A) Genetic association analysis results for the significant SNPs located within the *MIR29C* gene (left panel), linkage disequilibrium for the significant SNPs based on *LDmatrix* (middle left panel), GTEx eQTL results for *CD46* (middle right panel) and histone modification marks (right panel) for hippocampal cells. B) Correlation plots between CSF biomarkers for which SNPs located within *MIR29C* share common genetic association. *Legend*: Enhancer states: H3K27ac, H3K4me1; Promoter states: H3K4me3. Enh = Enhancer, EnhG: genic enhancer, TssAFlnk: flanking active TSS, TxFlnk: Transcr. At gene 5’ and 3’.

### Pathways enrichment analysis for the significant miRNAs

For the six miRNAs significantly dysregulated in the low performance group, we identified 1 641 experimentally validated miRNA-gene pairs in miRTarBase. Of these, 734 (44.71%) were highly expressed in the brain. MiR-128-3p targets the highest number of genes while hsa-miR-26a-5p targets the highest percentage of genes highly expressed in the brain (48.58%) (Figure 3B). Although there was no overlap between the targeted genes for the six miRNAs, 11 genes were targeted by at least three of the dysregulated miRNAs (Figure 3C). The pathway enrichment analysis conducted in KEGG, GO and REACTOME revealed 152 unique pathways (Figure 3A). Importantly, nine pathways were targeted by more than one miRNA; these pathways contribute to cell cycle regulation, transcription and splicing, cellular signalling and tyrosine kinase signalling. Moreover, the pathway cluster “Wnt/ β catenin signalling” was targeted by hsa-miR-26a-5p only, while pathways clusters related to inflammation such as “NF-κβ signalling”, “cytokine production” and “toll like receptor signalling” contained pathways were targeted by hsa-miR-146a-5p only (Supplementary material table 5).

**Figure 3.**
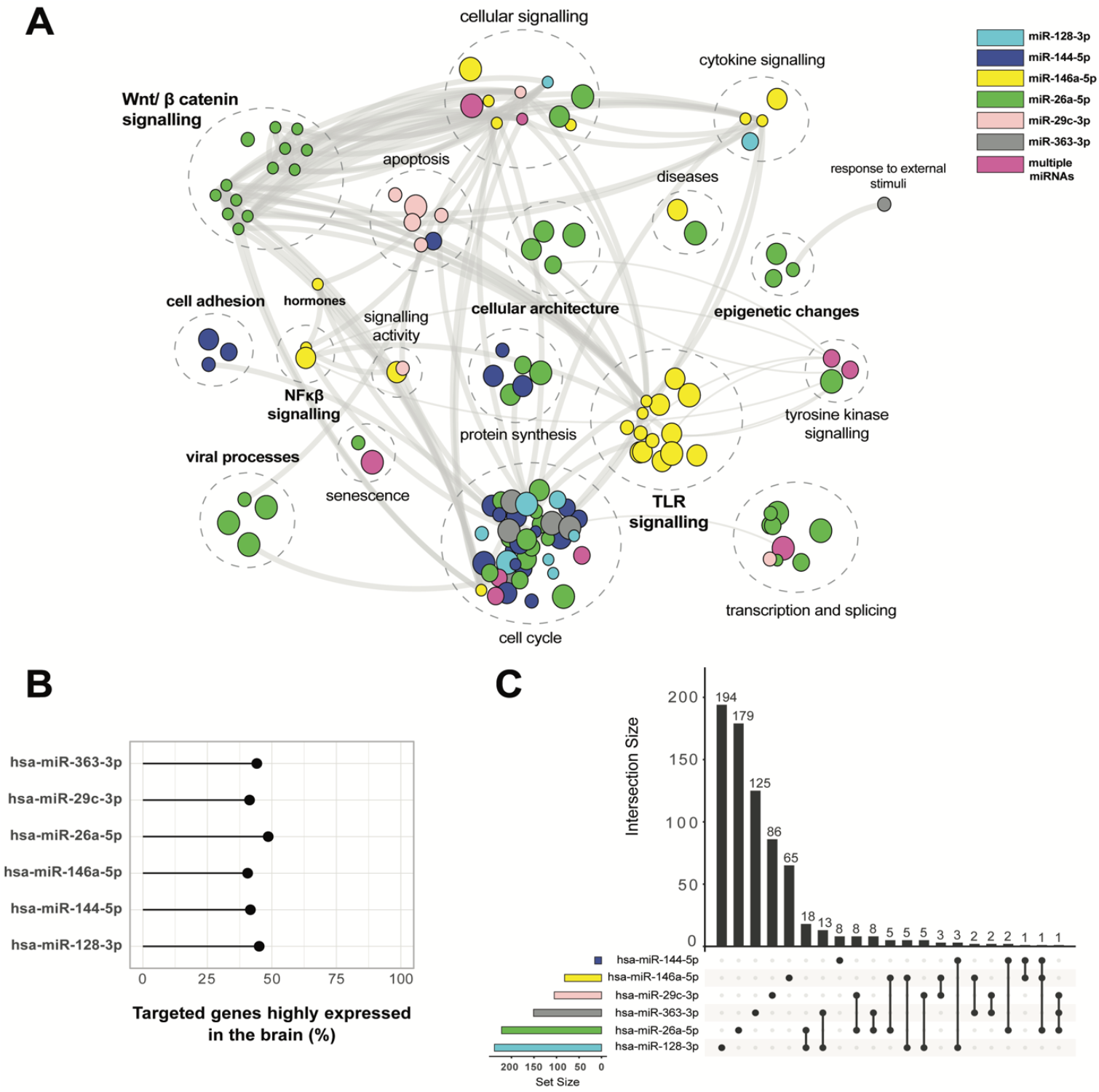
Functional enrichment for associated SNPs. A) Pathway enrichment analysis for the significant miRNAs differentially expressed between low and normal performers in CHARIOT PRO. In bold are clusters for which pathways are enriched by genes targeted by a single dysregulated miRNA. B) Percentage of targeted brain specific genes for the significant miRNAs C) Intersection between targeted brain specific genes for each significant miRNA.

### Functional enrichment for associated SNPs

Among *MIR29C* gene SNPs significantly associated with Aβ42, sTREM2 and BACE1 activity, nine variants are intronic of *CD34, CD46, LOC148696* genes. According to the VARAdb, these SNPs are associated with glioma, gene expression of *CD46, CD34* expression in dendritic cells, MIR29C as well as Parkinson’s disease (PD). Data from the Roadmap Epigenomics project reveals that these SNPs are in regulatory regions with enhancer/ promoter activity in hippocampal cells (Figure 2A). We did not identify SNPs located in DNase I hypersensitive sites. Of note, according to ChIP data from the ENCODE project, SNPs significantly associated with sTREM2 levels, BACE1 activity and Aβ42 levels alter the binding of several brain specific transcription factors (TF), critical in neurogenesis, microglial activation, apoptosis or neuronal response to Aβ disposition (Supplementary material table 6). Several SNPs associated with CSF BACE1 activity or Aβ42 levels are in LD with rs2724384 and rs4844390, two intronic variants of *CD46*. In this regard, analysis of eQTL data from the hippocampus in GTEx reveals that all SNPs associated with Aβ42 are eQTLs for *CD46*, suggesting that CD46 might mediate the effect of these SNPs on Aβ42. Additionally, eight (40%) of the 20 SNPs, significantly associated with BACE1 activity levels, were eQTLs for *CD46*; six reduced and two increased CD46 expression, respectively (Figure 2A). We did not find any significant eQTLs signals for *CD34, MIR29C* or *CR1*.

## Discussion

In this study, we dissected miRNA blood expression levels in 830 older individuals without MCI or dementia. We studied 38 miRNAs significantly associated with AD in our recent systematic literature review and meta-analysis ^14^. Six miRNAs (hsa-miR-128-3p, hsa-miR-144-5p, hsa-miR-146a-5p, hsa-miR-26a-5p, hsa-miR-29c-3p and hsa-miR-363-3p) were significantly downregulated in the blood of individuals with subtle cognitive deficit on the RBANS. This set of miRNAs could serve as predictive biomarker in individuals with pre-MCI cognitive deficits, since they may be dysregulated before the onset of overt clinical AD. To unveil the mechanistic role of the six-miRNAs signature, we undertook a genetic association analysis using 750 individuals from ADNI. Here, we found that SNPs within the *MIR29C* gene were significantly associated with Aβ42 and sTREM2 levels and BACE1 activity in the CSF. Our pathway enrichment analysis suggests a role for miR-29c in early phases of the disease. Supporting our results, a recent study in 76 subjects with MCI found that miR-146a-5p belonged to a set of three blood miRNAs (miR-181a-5p, miR-148a-3p and miR-146a-5p) predicting progression to AD dementia ^16^. The authors suggested that targeting this miRNA signature via RNA therapeutics ameliorates cognitive decline in an AD mouse model ^16^. MiR-181a-5p and miR-148a-3p were not associated with AD in our meta-analysis in blood, brain and CSF. However, the authors identified the target miRNAs in a cohort of healthy younger individuals; miR-181a-5p and miR-148a-3p may be differentially expressed several decades before disease onset, but normally expressed closer to the first symptoms ^16^.

Our in-silico pathway enrichment analysis showed that the set of six circulating miRNAs targets brain-specific genes involved in early AD pathological processes, including apoptosis and TLR and NF-κβ inflammatory signalling pathways. Strikingly, we found that genes from the Wnt/ β catenin system are targeted exclusively by miR-26a. Mounting evidence shows that this pathway plays a key role in AD: decreased levels of Wnt/ β catenin were associated with increased Aβ deposits and impaired cognitive function in an AD mouse model ^55, 56^. Moreover, β catenin also inhibited BACE1 expression leading to decreased Aβ formation ^57^. So far, the relationship between miR-26a and the Wnt/ β catenin system has not been explored in AD. In non-small cell lung cancer and cholangiocarcinoma, downregulation of miR-26a lead to lower β catenin levels ^58, 59^. Consequently, we could hypothesize that individuals in our cohort with lower cognitive performance and downregulated miR-26a had lower levels of Wnt and β catenin, resulting in decreased Aβ clearance. This hypothesis is of particular interest, since Wnt/ β catenin is targeted by statins, a class of drugs possibly reducing AD risk ^60^. In addition to miR-26a, we found that miR-146a targets brain specific genes involved in the NF-κβ and TLR signalling pathways. These two pathways are key actors in early AD pathological processes ^61, 62^. For instance, NF stimulates the expression of pro- and anti-inflammatory cytokines, such as IL1-β and TNF-α, before Aβ deposition ^63^.

So far, no other study has investigated the expression of miR-29c in a longitudinal cohort of patients from the AD continuum. Our pathway enrichment analysis suggests a role for miR-29c in early phases of the disease. Notably, we showed how miR-29c targets brain specific genes of the caspase family, playing a role early in AD ^64^. In line with our results, a recent study showed that administration of a caspase-1 inhibitor to pre-symptomatic APP mutant mice slowed down cognitive decline ^65^. Moreover, in a Parkinson’s disease mouse model, decreased intraneuronal α synuclein aggregation was reported following miR-29c induced inhibition of pro-inflammatory cytokines production and caspase-3 and caspase-9 expression ^66^. Therefore, early downregulation of miR-29c at preclinical stages of AD may promote the development of Aβ deposits by increasing the production of pro-inflammatory cytokines. A link between miR-29c and the amyloid cascade is supported by our genetic association findings, revealing an association between *MIR29C* polymorphisms and Aβ42 and BACE1 activity levels in the CSF. Indeed, carriers of 7 minor alleles showed lower Aβ42 and BACE1 activity levels suggesting a protective effect. These results confirm a previous study reporting a negative correlation between miR-29c expression and BACE1 and Aβ levels in 31 AD brains ^67^. Moreover, we found that some of these SNPs, including rs4844390, alter the binding of transcription factors involved in APP expression, such as CTCF.

We identified a positive correlation between BACE1 activity and sTREM2 levels in the CSF, suggesting an interplay between microglial activation and the amyloid cascade at early stages of the disease. To the best of our knowledge, this relationship has not been described so far in the literature. In individuals with MCI, both increased BACE1 activity and increased microglial activation have been reported on brain extracts and 11C-^®^PK11195 PET imaging ^68, 69^. Moreover, in an AD mouse model, early disease stages were characterised by microglial activation with increased expression of IL-1β and IL-6 cytokines as well as increased BACE1 activity ^70^. Nevertheless, the relationship between BACE1 and microglial activation needs to be explored further as one study reported an inverse relationship between sTREM2 and BACE1 activity: in an APP transgenic mouse, inhibiting BACE1 expression resulted in an increase in microglial activation ^71^. Our results suggest a complex interplay between BACE1 and microglial activation, and different stages of microglial activation within participants of the BACE1 inhibitor trials might have undermined the efficacy of the substances, contributing to the disappointing results ^72^.

We identified seven SNPs in *MIR29C* associated with Aβ and BACE1 activity levels as eQTLs for the increased or decreased expression of CD46, a protein in the complement cascade and substrate of the gamma-secretases Presenilin 1 and 2, participating in APP cleavage ^73^; hence, those SNPs’ effect on Aβ and BACE1 may be mediated by CD46. To date, there are no studies investigating the interaction between CD46 and the amyloid cascade. However, in 16HBE14o-cells, CD46 binds the herpesvirus HHV-6A resulting in increased TREM2 expression, Aβ42 production and microglial activation ^74^. Although we did not find any eQTL signals for SNPs associated with sTREM2, rs606714 affects the binding of transcription factors from the Forkhead box family, participating in microglial activation in neuronal cells ^75^. Further experimental studies are needed to explore the link between CD46 and Aβ deposition.

Some limitations of our study need to be mentioned. First, the population-based CHARIOT PRO cohort does not include neurodegenerative biomarker data and no long-term follow-up investigations. To offset those limitations, we used CSF results from the memory clinic-based ADNI cohort, including deep longitudinal clinical and biological phenotyping. Second, we were underpowered to investigate associations for less common SNPs, such as those located within the miRNA genes. Finally, the biological plausibility of circulating miRNAs associated with disease should be underpinned ideally by similar results for their expression in brain. Since no brain data was available for any of our cohorts, we performed a pathway enrichment analysis based on genes known to be highly expressed in the brain, including only evidence from experimental studies, and functional analysis of the significant SNPs only conducted for findings from hippocampal cells. Investigating dysregulation of a miRNA in the brain and the blood of the same patient is limited by methodological and ethical barriers ^76^. The recent development of miRNA-based imaging techniques for in vivo miRNA activity exploration could set the base for the development of novel miRNA tracers to be used in neuroimaging ^77^.

Overall, we provide initial experimental evidence on the diagnostic utility of a reproducible six-miRNAs signature associated with AD in a population of older individuals with subtle cognitive deficits, and provide mechanistic validation using CSF AD biomarkers. Our findings advocate the role of miRNAs as minimally invasive biomarker candidates for cognitive decline. Considering that the expression of the individual miRNAs was highly correlated, a single miRNA, such as miR-29c, could be used to flag individuals at risk for cognitive decline, followed by more detailed diagnostic assessments. This staged approach would result in a scalable, cost-effective approach with reduced burden on the affected individuals, healthcare systems and payers.

The major strength of our study is the careful combination of an evidence-based principled selection of the best miRNA candidates, followed by a validation in two large independent cohorts, providing information for different levels of cognitive deficits and on different levels of investigation (cognitive and biological). The development of a miRNA-based point of care diagnostic solution is urgently needed for the effective roll-out of upcoming disease modifying AD therapies.

## Data Availability

All data produced in the present work are contained in the manuscript

## Acknowledgements

We thank the CHARIOT PRO and ADNI participants and staff for their support, dedication and commitment. The authors thank Janssen Pharmaceuticals Research & Development, LLC. for funding CHARIOT PRO. The miRNA qPCR analysis was funded by Stevenage Bioscience Catalyst. For the ADNI, data collection and sharing for this project was funded by the Alzheimer’s Disease Neuroimaging Initiative (ADNI) (National Institutes of Health Grant U01 AG024904) and DOD ADNI (Department of Defense award number W81XWH-12-2-0012). ADNI is funded by the National Institute on Aging, the National Institute of Biomedical Imaging and Bioengineering, and through generous contributions of: AbbVie, Alzheimer’s Association; Alzheimer’s Drug Discovery Foundation; Araclon Biotech; BioClinica, Inc.; Biogen; Bristol-Myers Squibb Company; CereSpir, Inc.; Cogstate; Eisai Inc.; Elan Pharmaceuticals, Inc.; Eli Lilly and Company; EuroImmun; F. Hoffmann-La Roche Ltd and its affiliated company Genentech, Inc.; Fujirebio; GE Healthcare; IXICO Ltd.; Janssen Alzheimer Immunotherapy Research & Development, LLC.; Johnson & Johnson Pharmaceutical Research & Development LLC.; Lumosity; Lundbeck; Merck & Co., Inc.; Meso Scale Diagnostics, LLC.; NeuroRx Research; Neurotrack Technologies; Novartis Pharmaceuticals Corporation; Pfizer Inc.; Piramal Imaging; Servier; Takeda Pharmaceutical Company; and Transition Therapeutics. The Canadian Institutes of Health Research is providing funds to support ADNI clinical sites in Canada. Private sector contributions are facilitated by the Foundation for the National Institutes of Health (www.fnih.org). The grantee organization is the Northern California Institute for Research and Education, and the study is coordinated by the Alzheimer’s Therapeutic Research Institute at the University of Southern California. ADNI data are disseminated by the Laboratory for Neuro Imaging at the University of Southern California. A.S received funding from the Imperial College London President’s PhD Scholarship (2017-2021). I.P. is funded by the Diabetes UK (BDA number: 20/0006307), the European Union’s Horizon 2020 research and innovation programme (LONGITOOLS, H2020-SC1-2019-874739), Agence Nationale de la Recherche (PreciDIAB, ANR-18-IBHU-0001), by the European Union through the “Fonds européen de développement regional” (FEDER), by the “Conseil Régional des Hauts-de-France” (Hauts-de-France Regional Council) and by the “Métropole Européenne de Lille” (MEL, European Metropolis of Lille).

## Supplementary material

Supplementary information is available at MP’s website.

## Supplementary material description

Detailed description of qPCR analysis in CHARIOT PRO

Table 1: Percentage of NA (ie Ct values > 35) per marker; those in bold font were not included in the subsequent analyses

Table 2: Regression coefficient with RBANS as outcome variable and miRNA normalised Ct value as predictor, model adjusted for age, gender, education years, ethnicity, *APOE* ε4 carrier status

Table 3: Correlation matrix for the six significant miRNAs

Table 4: miRNA gene environment

Figure 1: haplotype blocks for *MIR29C*

Table 5: Pathway enrichment analysis of the six significantly dysregulated miRNAs in the blood for targeted genes highly expressed in the brain

Table 6: Role of selected brain specific transcription factors for which binding is affected by significant SNPs associated with Aβ42, BACE1 and sTREM2 levels in the CSF

## References

1. Patterson C. World alzheimer report 2018. 2018.

2. Cummings J. Lessons Learned from Alzheimer Disease: Clinical Trials with Negative Outcomes. Clinical and Translational Science 2018; 11(2): 147–152.

3. Cummings J, Lee G, Zhong K, Fonseca J, Taghva K. Alzheimer’s disease drug development pipeline: 2021. Alzheimer’s & Dementia: Translational Research & Clinical Interventions 2021; 7(1): e12179.

4. Sperling RA, Aisen PS, Beckett LA, Bennett DA, Craft S, Fagan AM et al. Toward defining the preclinical stages of Alzheimer’s disease: recommendations from the National Institute on Aging-Alzheimer’s Association workgroups on diagnostic guidelines for Alzheimer’s disease. Alzheimer’s & dementia : the journal of the Alzheimer’s Association 2011; 7(3): 280–292.

5. Jack Jr CR, Bennett DA, Blennow K, Carrillo MC, Dunn B, Haeberlein SB et al. NIA-AA Research Framework: Toward a biological definition of Alzheimer’s disease. Alzheimer’s & Dementia 2018; 14(4): 535–562.

6. Barthélemy NR, Li Y, Joseph-Mathurin N, Gordon BA, Hassenstab J, Benzinger TLS et al. A soluble phosphorylated tau signature links tau, amyloid and the evolution of stages of dominantly inherited Alzheimer’s disease. Nature Medicine 2020; 26(3): 398–407.

7. Suárez-Calvet M, Karikari TK, Ashton NJ, Lantero Rodríguez J, Milà-Alomà M, Gispert JD et al. Novel tau biomarkers phosphorylated at T181, T217 or T231 rise in the initial stages of the preclinical Alzheimer’s continuum when only subtle changes in Aβ pathology are detected. EMBO molecular medicine 2020; 12(12): e12921.

8. Wang L, Benzinger TL, Su Y, Christensen J, Friedrichsen K, Aldea P et al. Evaluation of Tau Imaging in Staging Alzheimer Disease and Revealing Interactions Between β-Amyloid and Tauopathy. JAMA neurology 2016; 73(9): 1070–1077.

9. Hanseeuw BJ, Betensky RA, Jacobs HIL, Schultz AP, Sepulcre J, Becker JA et al. Association of Amyloid and Tau With Cognition in Preclinical Alzheimer Disease: A Longitudinal Study. JAMA neurology 2019; 76(8): 915–924.

10. Karikari TK, Pascoal TA, Ashton NJ, Janelidze S, Benedet AL, Rodriguez JL et al. Blood phosphorylated tau 181 as a biomarker for Alzheimer’s disease: a diagnostic performance and prediction modelling study using data from four prospective cohorts. The Lancet Neurology 2020; 19(5): 422–433.

11. Janelidze S, Mattsson N, Palmqvist S, Smith R, Beach TG, Serrano GE et al. Plasma P-tau181 in Alzheimer’s disease: relationship to other biomarkers, differential diagnosis, neuropathology and longitudinal progression to Alzheimer’s dementia. Nat Med 2020; 26(3): 379–386.

12. Bartel DP. MicroRNAs: genomics, biogenesis, mechanism, and function. Cell 2004; 116(2): 281–297.

13. Dehghani R, Rahmani F, Rezaei N. MicroRNA in Alzheimer’s disease revisited: implications for major neuropathological mechanisms. Reviews in the Neurosciences 2018; 29(2): 161–182.

14. Takousis P, Sadlon A, Schulz J, Wohlers I, Dobricic V, Middleton L et al. Differential expression of microRNAs in Alzheimer’s disease brain, blood, and cerebrospinal fluid. Alzheimer’s & dementia : the journal of the Alzheimer’s Association 2019; 15(11): 1468–1477.

15. Lusardi TA, Phillips JI, Wiedrick JT, Harrington CA, Lind B, Lapidus JA et al. MicroRNAs in Human Cerebrospinal Fluid as Biomarkers for Alzheimer’s Disease. Journal of Alzheimer’s disease : JAD 2017; 55(3): 1223–1233.

16. Islam MR, Kaurani L, Berulava T, Heilbronner U, Budde M, Centeno TP et al. A microRNA signature that correlates with cognition and is a target against cognitive decline. EMBO molecular medicine 2021; 13(11): e13659.

17. Campos-Melo D, Droppelmann CA, He Z, Volkening K, Strong MJ. Altered microRNA expression profile in Amyotrophic Lateral Sclerosis: a role in the regulation of NFL mRNA levels. Molecular brain 2013; 6: 26.

18. Preische O, Schultz SA, Apel A, Kuhle J, Kaeser SA, Barro C et al. Serum neurofilament dynamics predicts neurodegeneration and clinical progression in presymptomatic Alzheimer’s disease. Nat Med 2019; 25(2): 277–283.

19. Jin Y, Tu Q, Liu M. MicroRNA-125b regulates Alzheimer’s disease through SphK1 regulation. Mol Med Rep 2018; 18(2): 2373–2380.

20. Couttas TA, Kain N, Daniels B, Lim XY, Shepherd C, Kril J et al. Loss of the neuroprotective factor Sphingosine 1-phosphate early in Alzheimer’s disease pathogenesis. Acta neuropathologica communications 2014; 2: 9.

21. Abelson JF, Kwan KY, O’Roak BJ, Baek DY, Stillman AA, Morgan TM et al. Sequence variants in SLITRK1 are associated with Tourette’s syndrome. Science (New York, NY) 2005; 310(5746): 317–320.

22. Moraghebi M, Maleki R, Ahmadi M, Negahi AA, Abbasi H, Mousavi P. In silico Analysis of Polymorphisms in microRNAs Deregulated in Alzheimer Disease. Frontiers in neuroscience 2021; 15.

23. Zhang B, Wang A, Xia C, Lin Q, Chen C. A single nucleotide polymorphism in primary-microRNA-146a reduces the expression of mature microRNA-146a in patients with Alzheimer’s disease and is associated with the pathogenesis of Alzheimer’s disease. Mol Med Rep 2015; 12(3): 4037–4042.

24. Liu S, Liu Y, Hao W, Wolf L, Kiliaan AJ, Penke B et al. TLR2 is a primary receptor for Alzheimer’s amyloid β peptide to trigger neuroinflammatory activation. Journal of immunology (Baltimore, Md : 1950) 2012; 188(3): 1098–1107.

25. Chopra N, Wang R, Maloney B, Nho K, Beck JS, Pourshafie N et al. MicroRNA-298 reduces levels of human amyloid-β precursor protein (APP), β-site APP-converting enzyme 1 (BACE1) and specific tau protein moieties. Molecular psychiatry 2021; 26(10): 5636–5657.

26. Udeh-Momoh CT, Watermeyer T, Price G, de Jager Loots CA, Reglinska-Matveyev N, Ropacki M et al. Protocol of the Cognitive Health in Ageing Register: Investigational, Observational and Trial Studies in Dementia Research (CHARIOT): Prospective Readiness cOhort (PRO) SubStudy. BMJ open 2021; 11(6): e043114.

27. Udeh-Momoh C, Price G, Ropacki MT, Ketter N, Andrews T, Arrighi HM et al. Prospective Evaluation of Cognitive Health and Related Factors in Elderly at Risk for Developing Alzheimer’s Dementia: A Longitudinal Cohort Study. The Journal of Prevention of Alzheimer’s Disease 2019; 6(4): 256–266.

28. Mueller SG, Weiner MW, Thal LJ, Petersen RC, Jack C, Jagust W et al. The Alzheimer’s Disease Neuroimaging Initiative. Neuroimaging Clinics of North America 2005; 15(4): 869–877.

29. McKhann G, Drachman D, Folstein M, Katzman R, Price D, Stadlan EM. Clinical diagnosis of Alzheimer’s disease: report of the NINCDS-ADRDA Work Group under the auspices of Department of Health and Human Services Task Force on Alzheimer’s Disease. Neurology 1984; 34(7): 939–944.

30. Petersen RC, Aisen PS, Beckett LA, Donohue MC, Gamst AC, Harvey DJ et al. Alzheimer’s Disease Neuroimaging Initiative (ADNI). Neurology 2010; 74(3): 201.

31. Randolph C, Tierney MC, Mohr E, Chase TN. The Repeatable Battery for the Assessment of Neuropsychological Status (RBANS): preliminary clinical validity. Journal of clinical and experimental neuropsychology 1998; 20(3): 310–319.

32. Randolph C. Rbans repeatable battery for the assessment of neuropsychological status: Manual. Psychological Corp.1999.

33. Karantzoulis S, Novitski J, Gold M, Randolph C. The Repeatable Battery for the Assessment of Neuropsychological Status (RBANS): Utility in detection and characterization of mild cognitive impairment due to Alzheimer’s disease. Archives of clinical neuropsychology : the official journal of the National Academy of Neuropsychologists 2013; 28(8): 837–844.

34. Duff K, Humphreys Clark JD, O’Bryant SE, Mold JW, Schiffer RB, Sutker PB. Utility of the RBANS in detecting cognitive impairment associated with Alzheimer’s disease: sensitivity, specificity, and positive and negative predictive powers. Archives of clinical neuropsychology : the official journal of the National Academy of Neuropsychologists 2008; 23(5): 603–612.

35. Zafari S, Backes C, Leidinger P, Meese E, Keller A. Regulatory MicroRNA Networks: Complex Patterns of Target Pathways for Disease-related and Housekeeping MicroRNAs. Genomics, Proteomics & Bioinformatics 2015; 13(3): 159–168.

36. Gevaert AB, Witvrouwen I, Vrints CJ, Heidbuchel H, Van Craenenbroeck EM, Van Laere SJ et al. MicroRNA profiling in plasma samples using qPCR arrays: Recommendations for correct analysis and interpretation. PLOS ONE 2018; 13(2): e0193173.

37. Barrett JC, Fry B, Maller J, Daly MJ. Haploview: analysis and visualization of LD and haplotype maps. Bioinformatics (Oxford, England) 2005; 21(2): 263–265.

38. Gabriel SB, Schaffner SF, Nguyen H, Moore JM, Roy J, Blumenstiel B et al. The structure of haplotype blocks in the human genome. Science (New York, NY) 2002; 296(5576): 2225–2229.

39. Saykin AJ, Shen L, Yao X, Kim S, Nho K, Risacher SL et al. Genetic studies of quantitative MCI and AD phenotypes in ADNI: Progress, opportunities, and plans. Alzheimer’s & dementia : the journal of the Alzheimer’s Association 2015; 11(7): 792–814.

40. Anderson CA, Pettersson FH, Clarke GM, Cardon LR, Morris AP, Zondervan KT. Data quality control in genetic case-control association studies. Nature protocols 2010; 5(9): 1564–1573.

41. Shaw LM, Vanderstichele H, Knapik-Czajka M, Clark CM, Aisen PS, Petersen RC et al. Cerebrospinal fluid biomarker signature in Alzheimer’s disease neuroimaging initiative subjects. Annals of neurology 2009; 65(4): 403–413.

42. Mattsson N, Andreasson U, Zetterberg H, Blennow K. Association of Plasma Neurofilament Light With Neurodegeneration in Patients With Alzheimer Disease. JAMA neurology 2017; 74(5): 557–566.

43. Kleinberger G, Yamanishi Y, Suárez-Calvet M, Czirr E, Lohmann E, Cuyvers E et al. TREM2 mutations implicated in neurodegeneration impair cell surface transport and phagocytosis. Science translational medicine 2014; 6(243): 243ra286–243ra286.

44. Suárez-Calvet M, Kleinberger G, Araque Caballero M, Brendel M, Rominger A, Alcolea D et al. sTREM2 cerebrospinal fluid levels are a potential biomarker for microglia activity in early-stage Alzheimer’s disease and associate with neuronal injury markers. EMBO molecular medicine 2016; 8(5): 466–476.

45. Wu G, Sankaranarayanan S, Tugusheva K, Kahana J, Seabrook G, Shi XP et al. Decrease in age-adjusted cerebrospinal fluid beta-secretase activity in Alzheimer’s subjects. Clinical biochemistry 2008; 41(12): 986–996.

46. Reimand J, Isserlin R, Voisin V, Kucera M, Tannus-Lopes C, Rostamianfar A et al. Pathway enrichment analysis and visualization of omics data using g:Profiler, GSEA, Cytoscape and EnrichmentMap. Nature protocols 2019; 14(2): 482–517.

47. Thul PJ, Lindskog C. The human protein atlas: A spatial map of the human proteome. Protein science : a publication of the Protein Society 2018; 27(1): 233–244.

48. Huang HY, Lin YC, Li J, Huang KY, Shrestha S, Hong HC et al. miRTarBase 2020: updates to the experimentally validated microRNA-target interaction database. Nucleic acids research 2020; 48(D1): D148–d154.

49. Conway JR, Lex A, Gehlenborg N. UpSetR: an R package for the visualization of intersecting sets and their properties. Bioinformatics (Oxford, England) 2017; 33(18): 2938–2940.

50. Machiela MJ, Chanock SJ. LDlink: a web-based application for exploring population-specific haplotype structure and linking correlated alleles of possible functional variants. Bioinformatics (Oxford, England) 2015; 31(21): 3555–3557.

51. Consortium; G. The Genotype-Tissue Expression (GTEx) project. Nature genetics 2013; 45(6): 580–585.

52. Ward LD, Kellis M. HaploReg: a resource for exploring chromatin states, conservation, and regulatory motif alterations within sets of genetically linked variants. Nucleic acids research 2012; 40(D1): D930–D934.

53. Ward LD, Kellis M. HaploReg v4: systematic mining of putative causal variants, cell types, regulators and target genes for human complex traits and disease. Nucleic acids research 2016; 44(D1): D877–881.

54. Pan Q, Liu Y-J, Bai X-F, Han X-L, Jiang Y, Ai B et al. VARAdb: a comprehensive variation annotation database for human. Nucleic acids research 2021; 49(D1): D1431–D1444.

55. Tapia-Rojas C, Inestrosa NC. Loss of canonical Wnt signaling is involved in the pathogenesis of Alzheimer’s disease. Neural regeneration research 2018; 13(10): 1705–1710.

56. De Ferrari GV, Avila ME, Medina MA, Perez-Palma E, Bustos BI, Alarcon MA. Wnt/β-catenin signaling in Alzheimer’s disease. CNS & neurological disorders drug targets 2014; 13(5): 745–754.

57. Parr C, Mirzaei N, Christian M, Sastre M. Activation of the Wnt/β-catenin pathway represses the transcription of the β-amyloid precursor protein cleaving enzyme (BACE1) via binding of T-cell factor-4 to BACE1 promoter. FASEB journal : official publication of the Federation of American Societies for Experimental Biology 2015; 29(2): 623–635.

58. Ye MF, Lin D, Li WJ, Xu HP, Zhang J. MiR-26a-5p Serves as an Oncogenic MicroRNA in Non-Small Cell Lung Cancer by Targeting FAF1. Cancer management and research 2020; 12: 7131–7142.

59. Zhang J, Han C, Wu T. MicroRNA-26a promotes cholangiocarcinoma growth by activating β-catenin. Gastroenterology 2012; 143(1): 246–256.e248.

60. McGuinness B, Craig D, Bullock R, Passmore P. Statins for the prevention of dementia. The Cochrane database of systematic reviews 2016; (1): Cd003160.

61. Momtazmanesh S, Perry G, Rezaei N. Toll-like receptors in Alzheimer’s disease. Journal of Neuroimmunology 2020; 348: 577362.

62. Ju Hwang C, Choi DY, Park MH, Hong JT. NF-κB as a Key Mediator of Brain Inflammation in Alzheimer’s Disease. CNS & neurological disorders drug targets 2019; 18(1): 3–10.

63. Cavanagh C, Colby-Milley J, Bouvier D, Farso M, Chabot J-G, Quirion R et al. βCTF-Correlated Burst of Hippocampal TNFα Occurs at a Very Early, Pre-Plaque Stage in the TgCRND8 Mouse Model of Alzheimer’s Disease. Journal of Alzheimer’s Disease 2013; 36: 233–238.

64. Flores J, Noël A, Foveau B, Lynham J, Lecrux C, LeBlanc AC. Caspase-1 inhibition alleviates cognitive impairment and neuropathology in an Alzheimer’s disease mouse model. Nature Communications 2018; 9(1): 3916.

65. Flores J, Noël A, Foveau B, Beauchet O, LeBlanc AC. Pre-symptomatic Caspase-1 inhibitor delays cognitive decline in a mouse model of Alzheimer disease and aging. Nature Communications 2020; 11(1): 4571.

66. Wang R, Yang Y, Wang H, He Y, Li C. MiR-29c protects against inflammation and apoptosis in Parkinson’s disease model in vivo and in vitro by targeting SP1. Clinical and Experimental Pharmacology and Physiology 2020; 47(3): 372–382.

67. Lei X, Lei L, Zhang Z, Zhang Z, Cheng Y. Downregulated miR-29c correlates with increased BACE1 expression in sporadic Alzheimer’s disease. International journal of clinical and experimental pathology 2015; 8(2): 1565–1574.

68. Cheng X, He P, Lee T, Yao H, Li R, Shen Y. High activities of BACE1 in brains with mild cognitive impairment. The American journal of pathology 2014; 184(1): 141–147.

69. Fan Z, Brooks DJ, Okello A, Edison P. An early and late peak in microglial activation in Alzheimer’s disease trajectory. Brain 2017; 140(3): 792–803.

70. Heneka MT, Sastre M, Dumitrescu-Ozimek L, Dewachter I, Walter J, Klockgether T et al. Focal glial activation coincides with increased BACE1 activation and precedes amyloid plaque deposition in APP[V717I] transgenic mice. Journal of neuroinflammation 2005; 2: 22.

71. Thakker DR, Sankaranarayanan S, Weatherspoon MR, Harrison J, Pierdomenico M, Heisel JM et al. Centrally Delivered BACE1 Inhibitor Activates Microglia, and Reverses Amyloid Pathology and Cognitive Deficit in Aged Tg2576 Mice. The Journal of neuroscience : the official journal of the Society for Neuroscience 2015; 35(17): 6931–6936.

72. Hampel H, Vassar R, De Strooper B, Hardy J, Willem M, Singh N et al. The β-Secretase BACE1 in Alzheimer’s Disease. Biological Psychiatry 2020.

73. Weyand NJ, Calton CM, Higashi DL, Kanack KJ, So M. Presenilin/gamma-secretase cleaves CD46 in response to Neisseria infection. Journal of immunology (Baltimore, Md : 1950) 2010; 184(2): 694–701.

74. Bortolotti D, Gentili V, Rotola A, Caselli E, Rizzo R. HHV-6A infection induces amyloid-beta expression and activation of microglial cells. Alzheimer’s Research & Therapy 2019; 11(1): 104.

75. Shang YC, Chong ZZ, Hou J, Maiese K. The forkhead transcription factor FOXO3a controls microglial inflammatory activation and eventual apoptotic injury through caspase 3. Current neurovascular research 2009; 6(1): 20–31.

76. Schott JM, Reiniger L, Thom M, Holton JL, Grieve J, Brandner S et al. Brain biopsy in dementia: clinical indications and diagnostic approach. Acta neuropathologica 2010; 120(3): 327–341.

77. Keshavarzi M, Sorayayi S, Jafar Rezaei M, Mohammadi M, Ghaderi A, Rostamzadeh A et al. MicroRNAs-Based Imaging Techniques in Cancer Diagnosis and Therapy. Journal of cellular biochemistry 2017; 118(12): 4121–4128.

