## Supplementary data for "A multi-omics approach identifies a blood-based miRNA signature of cognitive decline in two large observational trials"

**Detailed description of qPCR analysis in CHARIOT-PRO**

Briefly, RNA extraction was undertaken using the QIAsymphony PAXgene Blood RNA Kit on the QIAsymphony SP with 72 samples, in 3 batches of 24, per run, followed by spectrophotometry (Nanodrop) and gel densitometry (Agilent, TapeStation) for RNA quantification and quality control, respectively. Reverse transcription of 10 ng RNA per sample, in 10 μL reactions, was performed using the miRCURY LNA RT Kit (QIAGEN). cDNA was diluted 100x and assayed in 10 μL PCR reactions with the miRNA Ready-to-Use PCR Custom panel, using miRCURY LNA SYBR Green master mix, according to the miRCURY LNA miRNA PCR protocol; reactions, in 384-well plate format, were performed in a LightCycler 480 Real-Time PCR system (Roche), the amplification curves were visualised using the Roche LC software, Ct values determined, and provided to us for subsequent analyses

**Supplementary Table 1: Percentage of NA (ie Ct values > 35) per marker; those in bold font were not included in the subsequent analyses**

| **miRNA** | **percentage NA (%)** |
|---|---:|
| hsa-let-7a-5p | 3.75 |
| hsa-let-7c-5p | 0 |
| hsa-let-7d-3p | 0.94 |
| hsa-let-7d-5p | 0 |
| hsa-miR-107 | 0 |
| hsa-miR-125b-5p | 0.10 |
| hsa-miR-128-3p | 5.11 |
| hsa-miR-129-5p | 100 |
| hsa-miR-138-5p | 99.90 |
| hsa-miR-143-3p | 92.39 |
| hsa-miR-144-5p | 0.73 |
| hsa-miR-146a-5p | 1.77 |
| hsa-miR-150-3p | 92.28 |
| hsa-miR-15a-3p | 99.17 |
| hsa-miR-16-5p | 0 |
| hsa-miR-17-3p | 93.01 |
| hsa-miR-181c-5p | 98.64 |
| hsa-miR-191-5p | 0.21 |
| hsa-miR-195-5p | 99.48 |
| hsa-miR-19a-3p | 0.10 |
| hsa-miR-19a-5p | 99.79 |
| hsa-miR-21-3p | 94.99 |
| hsa-miR-210-3p | 3.34 |
| hsa-miR-26a-5p | 0 |
| hsa-miR-26b-3p | 4.80 |
| hsa-miR-27b-3p | 2.19 |
| hsa-miR-29c-3p | 2.61 |
| hsa-miR-30a-5p | 77.69 |
| hsa-miR-30d-5p | 0 |
| hsa-miR-31-5p | 78.42 |
| hsa-miR-340-3p | 62.57 |
| hsa-miR-342-3p | 0 |
| hsa-miR-361-5p | 0.31 |
| hsa-miR-363-3p | 0.21 |
| hsa-miR-425-5p | 0 |
| hsa-miR-454-3p | 1.36 |
| hsa-miR-455-5p | 98.33 |

| | |
|---|---|
| hsa-miR-483-3p | 86.34 |
| hsa-miR-5001-3p | 69.97 |
| hsa-miR-501-3p | 3.75 |
| hsa-miR-550a-3p | 0.10 |
| hsa-miR-671-3p | 71.95 |
| hsa-miR-885-5p | 96.25 |
| hsa-miR-92a-3p | 0 |
| hsa-miR-93-5p | 0 |
| hsa-miR-98-5p | 26.38 |
| UniSp3 | 0.31 |
| UniSp6 | 0 |

**Legend:** removed markers are in bold

**Supplementary Table 2: Regression coefficient with RBANS as outcome variable and miRNA normalised Ct value as predictor, model adjusted for age, gender, education years, ethnicity, APOE ε4 carrier status**

| domain | miRNA | b | [95% CI] | Std. Error | t value | Pr(>\|t\|) | FDR adj. P value |
|---|---|---|---|---|---|---|---|
| **Language Index** | hsa.let.7a.5p | -2.39 | [-4.10, -0.69] | 0.868 | -2.752 | 0.006 | 0.048 |
| | hsa.let.7c.5p | -2.26 | [-3.97, -0.56] | 0.871 | -2.601 | 0.009 | 0.049 |
| | hsa.let.7d.5p | -3.06 | [-1.67, 1.75] | 1.023 | -2.992 | 0.003 | 0.048 |
| | hsa.miR.144.5p | -0.89 | [-1.53, -0.25] | 0.327 | -2.723 | 0.007 | 0.048 |
| | hsa.miR.93.5p | -1.88 | [-3.31, -0.45] | 0.728 | -2.577 | 0.01 | 0.049 |
| | hsa.miR.98.5p | -1.95 | [-3.34, -0.55] | 0.712 | -2.735 | 0.006 | 0.048 |
| **Attention Index** | hsa.miR.363.3p | -4.27 | [-6.70, -1.85] | 1.236 | -3.457 | 0.001 | 0.017 |
| **Total Scale** | hsa.miR.144.5p | -1.17 | [-1.84, -0.51] | 0.339 | -3.464 | 0.001 | 0.016 |
| Ethnicity coded as white = 0, other = 1; APOE genotype coded as e3/e4 or e4/e4 = 1, other = 0 | | | | | | | |

**Supplementary Table 3: Correlation matrix for the six significant miRNAs**

**rho (spearman)**

| | miR.128.3p | miR.144.5p | miR.146a.5p | miR.26a.5p | miR.29c.3p | miR.363.3p |
|---|---|---|---|---|---|---|
| miR.128.3p | | | | | | |
| miR.144.5p | 0.09 | | | | | |
| miR.146a.5p | 0.27 | 0.38 | | | | |
| miR.26a.5p | 0.33 | 0.43 | 0.42 | | | |
| miR.29c.3p | 0.15 | 0.45 | 0.46 | 0.41 | | |
| miR.363.3p | 0.26 | 0.35 | 0.46 | 0.52 | 0.55 | |

**N**

| | miR.128.3p | miR.144.5p | miR.146a.5p | miR.26a.5p | miR.29c.3p | miR.363.3p |
|---|---|---|---|---|---|---|
| miR.128.3p | | | | | | |
| miR.144.5p | 798 | | | | | |
| miR.146a.5p | 804 | 796 | | | | |
| miR.26a.5p | 805 | 802 | 801 | | | |
| miR.29c.3p | 799 | 795 | 798 | 798 | | |
| miR.363.3p | 804 | 798 | 801 | 804 | 798 | |

**P values**

| | miR.128.3p | miR.144.5p | miR.146a.5p | miR.26a.5p | miR.29c.3p | miR.363.3p |
|---|---|---|---|---|---|---|
| miR.128.3p | | | | | | |
| miR.144.5p | 0.0128 | | | | | |
| miR.146a.5p | <0.0001 | <0.0001 | | | | |
| miR.26a.5p | <0.0001 | <0.0001 | <0.0001 | | | |
| miR.29c.3p | <0.0001 | <0.0001 | <0.0001 | <0.0001 | | |
| miR.363.3p | <0.0001 | <0.0001 | <0.0001 | <0.0001 | <0.0001 | |

**Supplementary Table 4: miRNA gene environment**

| RefSeq | chr | strand | length | start | end | N SNPs* | RefSeq of genes within ± 200 kb of the gene |
|---|---|---|---|---|---|---|---|
| *MIR128-1* | 2 | + | 82 | 136422967 | 136423048 | 60 | *R3HDM1, MIR128-1* |
| *MIR144* | 17 | - | 86 | 27188551 | 27188636 | 99 | *SDF2, SUPT6H, PROCA1, RAB34, RPL23A, SNORD42B, SNORD4A, SNORD42B, SNORD4B, SNORD42A, TLCD1, NEK8, TRAF4, FAM222B, ERAL1, MIR451A, MIR451B, MIR144, MIR4732, FLOT2, DHRS13, PHF12, LOC101927018, SEZ6, PIPOX* |
| *MIR146A* | 5 | + | 99 | 159912359 | 159912457 | 206 | *MIR3142HG, MIR146A* |
| *MIR26A1* | 3 | + | 77 | 38010895 | 38010971 | 109 | *CTDSPL, MIR26A1* |
| *MIR29C* | 1 | - | 88 | 207975197 | 207975284 | 181 | *CR1, CR1L, CD46, MIR29B2CHG, MIR29C, MIR29B2, LOC148696, CD34* |
| *MIR363* | X | - | 75 | 133303408 | 133303482 | 25 | *MIR363* |

*number of SNPS Minor Allele Frequency $\geq 0.05$
Legend: chr: chromosome, RefSeq National Center for Biotechnology Information Reference Sequence, SNP: Single Nucleotide Polymorphism

**Supplementary figure 1: haplotype blocks MIR29C**

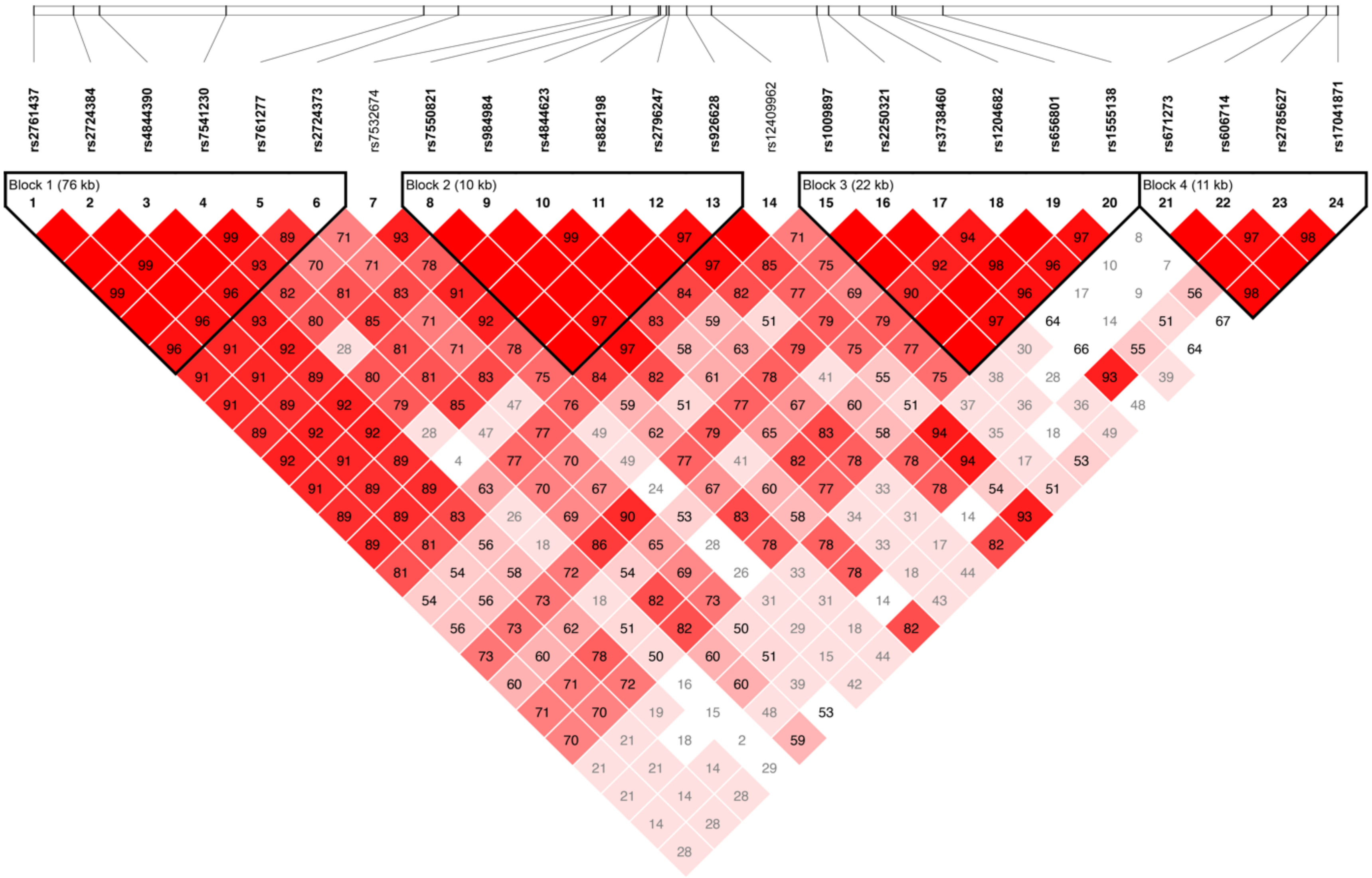

**Supplementary Table 5: Pathway enrichment analysis of the six significantly dysregulated miRNAs in the blood for targeted genes highly expressed in the brain**

| ID | Description | FDR | intersection | cluster | mirnas | N mirnas |
|---|---|---|---|---|---|---|
| GO:1905710 | positive regulation of membrane permeability | 0.005 | *GSK3B,SLC25A5,YWHAE,BLOC1S2* | cellular architecture | hsa-miR-26a-5p | 1 |
| REAC:R-HSA-975110 | TRAF6 mediated IRF7 activation in TLR7/8 or 9 signaling | 0.006 | *IRAK1,IRF7* | toll like receptor signaling pathway | hsa-miR-146a-5p | 1 |
| GO:0006476 | protein deacetylation | 0.006 | *RCOR1,PHB,SFPQ,MTA3* | protein synthesis | hsa-miR-26a-5p | 1 |
| GO:0090559 | regulation of membrane permeability | 0.006 | *GSK3B,SLC25A5,YWHAE,BLOC1S2* | cellular architecture | hsa-miR-26a-5p | 1 |
| REAC:R-HSA-9006925 | Intracellular signaling by second messengers | 0.007 | *GSK3B,RPS27A,TNRC6B,RCOR1,MDM2,PPP2R5D,MTA3,PRKX* | cellular signaling | hsa-miR-146a-5p, hsa-miR-146a-5p, hsa-miR-26a-5p, hsa-miR-26a-5p | 4 |
| REAC:R-HSA-9006925 | Intracellular signaling by second messengers | 0.007 | *GSK3B,RPS27A,TNRC6B,RCOR1,MDM2,PPP2R5D,MTA3,PRKX* | cellular signaling | hsa-miR-146a-5p, hsa-miR-146a-5p, hsa-miR-26a-5p, hsa-miR-26a-5p | 4 |
| GO:0098732 | macromolecule deacylation | 0.007 | *RCOR1,PHB,SFPQ,MTA3* | protein synthesis | hsa-miR-26a-5p | 1 |
| GO:1905214 | regulation of RNA binding | 0.009 | *CDK9,NUCKS1* | transcription and splicing | hsa-miR-26a-5p | 1 |
| REAC:R-HSA-1257604 | PIP3 activates AKT signaling | 0.009 | *GSK3B,RPS27A,TNRC6B,RCOR1,MDM2,PPP2R5D,MTA3* | tyrosine kinase signaling | hsa-miR-26a-5p | 1 |
| REAC:R-HSA-212165 | Epigenetic regulation of gene expression | 0.009 | *GSK3B,UBTF,TDG,MTA3,POLR2E* | epigenetic changes | hsa-miR-26a-5p | 1 |
| REAC:R-HSA-3700989 | Transcriptional Regulation by TP53 | 0.009 | *CCNE1,RPS27A,YWHAE,TNRC6B,MDM2,CDK9,POLR2E,COX5A* | transcription and splicing | hsa-miR-26a-5p, hsa-miR-29c-3p | 2 |
| GO:0016032 | viral process | 0.013 | *RPS27A,DDB1,PDE12,PHB,CDK9,POLR2E,NUCKS1* | viral processes | hsa-miR-26a-5p | 1 |
| GO:0031570 | DNA integrity checkpoint | 0.014 | *MDM2,CNOT4,CDC5L,CNOT2* | cell cycle | hsa-miR-363-3p | 1 |
| GO:0044783 | G1 DNA damage checkpoint | 0.015 | *MDM2,CNOT4,CNOT2* | cell cycle | hsa-miR-363-3p | 1 |
| REAC:R-HSA-198323 | AKT phosphorylates targets in the cytosol | 0.017 | *AKT3,MDM2* | cellular signaling | hsa-miR-26a-5p, hsa-miR-29c-3p | 2 |
| REAC:R-HSA-6804759 | Regulation of TP53 Activity through Association with Co-factors | 0.017 | *PPP1R13B,AKT3* | cellular signaling | hsa-miR-29c-3p | 1 |
| GO:0006283 | transcription-coupled nucleotide-excision repair | 0.021 | *RPS27A,DDB1,POLR2E* | cell cycle | hsa-miR-26a-5p | 1 |
| GO:0090305 | nucleic acid phosphodiester bond hydrolysis | 0.021 | *RPS27A,DDB1,PDE12,CPSF2* | cell cycle | hsa-miR-26a-5p | 1 |
| GO:0019058 | viral life cycle | 0.023 | *RPS27A,DDB1,PDE12,PHB,NUCKS1* | viral processes | hsa-miR-26a-5p | 1 |
| REAC:R-HSA-8849470 | PTK6 Regulates Cell Cycle | 0.024 | *CCNE1* | cell cycle | hsa-miR-144-5p | 1 |
| GO:0046677 | response to antibiotic | 0.026 | *MDM2,RPL23* | response to external stimuli | hsa-miR-363-3p | 1 |

| | | | | | | |
|---|---|---|---|---|---|---|
| REAC:R-HSA-166166 | MyD88-independent TLR4 cascade | 0.026 | *IRAK1,IRF7* | toll like receptor signaling pathway | hsa-miR-146a-5p | 1 |
| REAC:R-HSA-168138 | Toll Like Receptor 9 (TLR9) Cascade | 0.026 | *IRAK1,IRF7* | toll like receptor signaling pathway | hsa-miR-146a-5p | 1 |
| REAC:R-HSA-168164 | Toll Like Receptor 3 (TLR3) Cascade | 0.026 | *IRAK1,IRF7* | toll like receptor signaling pathway | hsa-miR-146a-5p | 1 |
| REAC:R-HSA-168181 | Toll Like Receptor 7/8 (TLR7/8) Cascade | 0.026 | *IRAK1,IRF7* | toll like receptor signaling pathway | hsa-miR-146a-5p | 1 |
| REAC:R-HSA-9006925 | Intracellular signaling by second messengers | 0.026 | *IRAK1,MTA2,PRKCE* | cellular signaling | hsa-miR-146a-5p, hsa-miR-146a-5p, hsa-miR-26a-5p, hsa-miR-26a-5p | 4 |
| REAC:R-HSA-9006925 | Intracellular signaling by second messengers | 0.026 | *IRAK1,MTA2,PRKCE* | cellular signaling | hsa-miR-146a-5p, hsa-miR-146a-5p, hsa-miR-26a-5p, hsa-miR-26a-5p | 4 |
| REAC:R-HSA-937061 | TRIF(TICAM1)-mediated TLR4 signaling | 0.026 | *IRAK1,IRF7* | toll like receptor signaling pathway | hsa-miR-146a-5p | 1 |
| GO:0000956 | nuclear-transcribed mRNA catabolic process | 0.027 | *CNOT4,CNOT2,RPL23,RPL24* | cell cycle | hsa-miR-363-3p | 1 |
| GO:0002221 | pattern recognition receptor signaling pathway | 0.029 | *IRAK1,IRF7,PRKCE* | cellular signaling | hsa-miR-146a-5p | 1 |
| GO:0002224 | toll-like receptor signaling pathway | 0.029 | *IRAK1,IRF7,PRKCE* | toll like receptor signaling pathway | hsa-miR-146a-5p | 1 |
| GO:0002755 | MyD88-dependent toll-like receptor signaling pathway | 0.029 | *IRAK1,IRF7* | toll like receptor signaling pathway | hsa-miR-146a-5p | 1 |
| GO:0002756 | MyD88-independent toll-like receptor signaling pathway | 0.029 | *IRF7,PRKCE* | toll like receptor signaling pathway | hsa-miR-146a-5p | 1 |
| GO:0034142 | toll-like receptor 4 signaling pathway | 0.029 | *IRAK1,PRKCE* | toll like receptor signaling pathway | hsa-miR-146a-5p | 1 |
| REAC:R-HSA-166016 | Toll Like Receptor 4 (TLR4) Cascade | 0.03 | *IRAK1,IRF7* | toll like receptor signaling pathway | hsa-miR-146a-5p | 1 |
| GO:0051702 | biological process involved in interaction with symbiont | 0.031 | *DDB1,PHB,NUCKS1* | cellular signaling | hsa-miR-26a-5p | 1 |
| REAC:R-HSA-110357 | Displacement of DNA glycosylase by APEX1 | 0.033 | *MBD4* | cell cycle | hsa-miR-146a-5p | 1 |
| REAC:R-HSA-168898 | Toll-like Receptor Cascades | 0.033 | *IRAK1,IRF7* | toll like receptor signaling pathway | hsa-miR-146a-5p | 1 |
| REAC:R-HSA-3134963 | DEx/H-box helicases activate type I IFN and inflammatory cytokines production | 0.033 | *IRF7* | cytokine | hsa-miR-146a-5p | 1 |
| REAC:R-HSA-3304351 | Signaling by TGF-beta Receptor Complex in Cancer | 0.033 | *SMAD2* | cytokine | hsa-miR-146a-5p | 1 |
| REAC:R-HSA-2559585 | Oncogene Induced Senescence | 0.033 | *RPS27A,TNRC6B,MDM2* | senescence | hsa-miR-26a-5p | 1 |
| GO:0019080 | viral gene expression | 0.033 | *RPS27A,CDK9,POLR2E,NUCKS1* | viral processes | hsa-miR-26a-5p | 1 |
| GO:0070911 | global genome nucleotide-excision repair | 0.033 | *RPS27A,DDB1* | cell cycle | hsa-miR-26a-5p | 1 |
| REAC:R-HSA-114452 | Activation of BH3-only proteins | 0.034 | *PPP1R13B,AKT3* | apoptosis and senescence | hsa-miR-29c-3p | 1 |
| REAC:R-HSA-2173796 | SMAD2/SMAD3:SMAD4 heterotrimer regulates transcription | 0.034 | *CCNT2,WWTR1* | transcription and splicing | hsa-miR-29c-3p | 1 |

| | | | | | | |
|---|---|---|---|---|---|---|
| REAC:R-HSA-3700989 | Transcriptional Regulation by TP53 | 0.034 | *PPP1R13B,AKT3,CCNT2,MDM2* | transcription and splicing | hsa-miR-26a-5p, hsa-miR-29c-3p | 2 |
| REAC:R-HSA-5633007 | Regulation of TP53 Activity | 0.034 | *PPP1R13B,AKT3,MDM2* | apoptosis and senescence | hsa-miR-29c-3p | 1 |
| REAC:R-HSA-5674400 | Constitutive Signaling by AKT1 E17K in Cancer | 0.034 | *AKT3,MDM2* | signaling activity | hsa-miR-29c-3p | 1 |
| REAC:R-HSA-6804757 | Regulation of TP53 Degradation | 0.034 | *AKT3,MDM2* | apoptosis and senescence | hsa-miR-29c-3p | 1 |
| REAC:R-HSA-6806003 | Regulation of TP53 Expression and Degradation | 0.034 | *AKT3,MDM2* | apoptosis and senescence | hsa-miR-29c-3p | 1 |
| REAC:R-HSA-1538133 | G0 and Early G1 | 0.034 | *CCNE1* | cell cycle | hsa-miR-144-5p | 1 |
| REAC:R-HSA-1638091 | Heparan sulfate/heparin (HS-GAG) metabolism | 0.034 | *HS3ST1* | hormone and metabolites | hsa-miR-144-5p | 1 |
| REAC:R-HSA-2022928 | HS-GAG biosynthesis | 0.034 | *HS3ST1* | hormone and metabolites | hsa-miR-144-5p | 1 |
| REAC:R-HSA-2559586 | DNA Damage/Telomere Stress Induced Senescence | 0.034 | *CCNE1* | apoptosis and senescence | hsa-miR-144-5p | 1 |
| REAC:R-HSA-390471 | Association of TriC/CCT with target proteins during biosynthesis | 0.034 | *CCNE1* | protein synthesis | hsa-miR-144-5p | 1 |
| REAC:R-HSA-6791312 | TP53 Regulates Transcription of Cell Cycle Genes | 0.034 | *CCNE1* | cell cycle | hsa-miR-144-5p | 1 |
| REAC:R-HSA-6804116 | TP53 Regulates Transcription of Genes Involved in G1 Cell Cycle Arrest | 0.034 | *CCNE1* | cell cycle | hsa-miR-144-5p | 1 |
| REAC:R-HSA-69017 | CDK-mediated phosphorylation and removal of Cdc6 | 0.034 | *CCNE1* | cell cycle | hsa-miR-144-5p | 1 |
| REAC:R-HSA-69205 | G1/S-Specific Transcription | 0.034 | *CCNE1* | cell cycle | hsa-miR-144-5p | 1 |
| REAC:R-HSA-69563 | p53-Dependent G1 DNA Damage Response | 0.034 | *CCNE1* | cell cycle | hsa-miR-144-5p, hsa-miR-144-5p, hsa-miR-26a-5p, hsa-miR-26a-5p | 4 |
| REAC:R-HSA-69563 | p53-Dependent G1 DNA Damage Response | 0.034 | *CCNE1* | cell cycle | hsa-miR-144-5p, hsa-miR-144-5p, hsa-miR-26a-5p, hsa-miR-26a-5p | 4 |
| REAC:R-HSA-69580 | p53-Dependent G1/S DNA damage checkpoint | 0.034 | *CCNE1* | cell cycle | hsa-miR-144-5p, hsa-miR-144-5p, hsa-miR-26a-5p, hsa-miR-26a-5p | 4 |
| REAC:R-HSA-69580 | p53-Dependent G1/S DNA damage checkpoint | 0.034 | *CCNE1* | cell cycle | hsa-miR-144-5p, hsa-miR-144-5p, hsa-miR-26a-5p, hsa-miR-26a-5p | 4 |
| REAC:R-HSA-69615 | G1/S DNA Damage Checkpoints | 0.034 | *CCNE1* | cell cycle | hsa-miR-144-5p, hsa-miR-144-5p, hsa-miR-26a-5p, hsa-miR-26a-5p | 4 |
| REAC:R-HSA-69615 | G1/S DNA Damage Checkpoints | 0.034 | *CCNE1* | cell cycle | hsa-miR-144-5p, hsa-miR-144-5p, hsa-miR-26a-5p, hsa-miR-26a-5p | 4 |
| REAC:R-HSA-8848021 | Signaling by PTK6 | 0.034 | *CCNE1* | tyrosine kinase signaling | hsa-miR-144-5p, hsa-miR-144-5p, hsa-miR-26a-5p, hsa-miR-26a-5p | 4 |

| | | | | | | |
|---|---|---|---|---|---|---|
| REAC:R-HSA-8848021 | Signaling by PTK6 | 0.034 | *CCNE1* | tyrosine kinase signaling | hsa-miR-144-5p, hsa-miR-144-5p, hsa-miR-26a-5p, hsa-miR-26a-5p | 4 |
| REAC:R-HSA-9006927 | Signaling by Non-Receptor Tyrosine Kinases | 0.034 | *CCNE1* | tyrosine kinase signaling | hsa-miR-144-5p, hsa-miR-144-5p, hsa-miR-26a-5p, hsa-miR-26a-5p | 4 |
| REAC:R-HSA-9006927 | Signaling by Non-Receptor Tyrosine Kinases | 0.034 | *CCNE1* | tyrosine kinase signaling | hsa-miR-144-5p, hsa-miR-144-5p, hsa-miR-26a-5p, hsa-miR-26a-5p | 4 |
| GO:0016311 | dephosphorylation | 0.036 | *GSK3B,YWHAE,PTPN13,PPP2R5D,MTMR12* | cellular signaling | hsa-miR-26a-5p | 1 |
| REAC:R-HSA-69202 | Cyclin E associated events during G1/S transition | 0.036 | *CCNE1* | cell cycle | hsa-miR-144-5p | 1 |
| REAC:R-HSA-69656 | Cyclin A:Cdk2-associated events at S phase entry | 0.036 | *CCNE1* | cell cycle | hsa-miR-144-5p | 1 |
| REAC:R-HSA-69052 | Switching of origins to a post-replicative state | 0.037 | *CCNE1* | cell cycle | hsa-miR-144-5p | 1 |
| REAC:R-HSA-390466 | Chaperonin-mediated protein folding | 0.037 | *CCNE1* | protein synthesis | hsa-miR-144-5p | 1 |
| REAC:R-HSA-112382 | Formation of RNA Pol II elongation complex | 0.037 | *RTF1,CDK9,POLR2E* | transcription and splicing | hsa-miR-26a-5p | 1 |
| REAC:R-HSA-198323 | AKT phosphorylates targets in the cytosol | 0.037 | *GSK3B,MDM2* | cellular signaling | hsa-miR-26a-5p, hsa-miR-29c-3p | 2 |
| REAC:R-HSA-4839735 | Signaling by AXIN mutants | 0.037 | *GSK3B,PPP2R5D* | Wnt/beta-catenin signaling pathway | hsa-miR-26a-5p | 1 |
| REAC:R-HSA-4839743 | Signaling by CTNNB1 phospho-site mutants | 0.037 | *GSK3B,PPP2R5D* | Wnt/beta-catenin signaling pathway | hsa-miR-26a-5p | 1 |
| REAC:R-HSA-4839744 | Signaling by APC mutants | 0.037 | *GSK3B,PPP2R5D* | Wnt/beta-catenin signaling pathway | hsa-miR-26a-5p | 1 |
| REAC:R-HSA-4839748 | Signaling by AMER1 mutants | 0.037 | *GSK3B,PPP2R5D* | Wnt/beta-catenin signaling pathway | hsa-miR-26a-5p | 1 |
| REAC:R-HSA-5339716 | Signaling by GSK3beta mutants | 0.037 | *GSK3B,PPP2R5D* | Wnt/beta-catenin signaling pathway | hsa-miR-26a-5p | 1 |
| REAC:R-HSA-5358747 | S33 mutants of beta-catenin aren't phosphorylated | 0.037 | *GSK3B,PPP2R5D* | Wnt/beta-catenin signaling pathway | hsa-miR-26a-5p | 1 |
| REAC:R-HSA-5358749 | S37 mutants of beta-catenin aren't phosphorylated | 0.037 | *GSK3B,PPP2R5D* | Wnt/beta-catenin signaling pathway | hsa-miR-26a-5p | 1 |
| REAC:R-HSA-5358751 | S45 mutants of beta-catenin aren't phosphorylated | 0.037 | *GSK3B,PPP2R5D* | Wnt/beta-catenin signaling pathway | hsa-miR-26a-5p | 1 |
| REAC:R-HSA-5358752 | T41 mutants of beta-catenin aren't phosphorylated | 0.037 | *GSK3B,PPP2R5D* | Wnt/beta-catenin signaling pathway | hsa-miR-26a-5p | 1 |
| REAC:R-HSA-5467337 | APC truncation mutants have impaired AXIN binding | 0.037 | *GSK3B,PPP2R5D* | Wnt/beta-catenin signaling pathway | hsa-miR-26a-5p | 1 |
| REAC:R-HSA-5467340 | AXIN missense mutants destabilize the destruction complex | 0.037 | *GSK3B,PPP2R5D* | Wnt/beta-catenin signaling pathway | hsa-miR-26a-5p | 1 |
| REAC:R-HSA-5467348 | Truncations of AMER1 destabilize the destruction complex | 0.037 | *GSK3B,PPP2R5D* | Wnt/beta-catenin signaling pathway | hsa-miR-26a-5p | 1 |
| REAC:R-HSA-6781823 | Formation of TC-NER Pre-Incision Complex | 0.037 | *RPS27A,DDB1,POLR2E* | cell cycle | hsa-miR-26a-5p | 1 |
| REAC:R-HSA-73762 | RNA Polymerase I Transcription Initiation | 0.037 | *UBTF,MTA3,POLR2E* | transcription and splicing | hsa-miR-26a-5p | 1 |

| | | | | | | |
|---|---|---|---|---|---|---|
| REAC:R-HSA-75955 | RNA Polymerase II Transcription Elongation | 0.037 | *RTF1,CDK9,POLR2E* | transcription and splicing | hsa-miR-26a-5p | 1 |
| REAC:R-HSA-8848021 | Signaling by PTK6 | 0.037 | *CCNE1,RPS27A,SFPQ* | tyrosine kinase signaling | hsa-miR-144-5p, hsa-miR-144-5p, hsa-miR-26a-5p, hsa-miR-26a-5p | 4 |
| REAC:R-HSA-8848021 | Signaling by PTK6 | 0.037 | *CCNE1,RPS27A,SFPQ* | tyrosine kinase signaling | hsa-miR-144-5p, hsa-miR-144-5p, hsa-miR-26a-5p, hsa-miR-26a-5p | 4 |
| REAC:R-HSA-9006927 | Signaling by Non-Receptor Tyrosine Kinases | 0.037 | *CCNE1,RPS27A,SFPQ* | tyrosine kinase signaling | hsa-miR-144-5p, hsa-miR-144-5p, hsa-miR-26a-5p, hsa-miR-26a-5p | 4 |
| REAC:R-HSA-9006927 | Signaling by Non-Receptor Tyrosine Kinases | 0.037 | *CCNE1,RPS27A,SFPQ* | tyrosine kinase signaling | hsa-miR-144-5p, hsa-miR-144-5p, hsa-miR-26a-5p, hsa-miR-26a-5p | 4 |
| REAC:R-HSA-391251 | Protein folding | 0.037 | *CCNE1* | protein synthesis | hsa-miR-144-5p | 1 |
| REAC:R-HSA-2468052 | Establishment of Sister Chromatid Cohesion | 0.038 | *STAG1,PDS5A* | cell cycle | hsa-miR-128-3p | 1 |
| REAC:R-HSA-2470946 | Cohesin Loading onto Chromatin | 0.038 | *STAG1,PDS5A* | cell cycle | hsa-miR-128-3p | 1 |
| REAC:R-HSA-2555396 | Mitotic Metaphase and Anaphase | 0.038 | *STAG1,LMNB1,KPNB1,PDS5A,TNPO1* | cell cycle | hsa-miR-128-3p | 1 |
| REAC:R-HSA-447115 | Interleukin-12 family signaling | 0.038 | *LMNB1,HNRNPF,CANX* | cytokine | hsa-miR-128-3p | 1 |
| REAC:R-HSA-68882 | Mitotic Anaphase | 0.038 | *STAG1,LMNB1,KPNB1,PDS5A,TNPO1* | cell cycle | hsa-miR-128-3p | 1 |
| REAC:R-HSA-68884 | Mitotic Telophase/Cytokinesis | 0.038 | *STAG1,PDS5A* | cell cycle | hsa-miR-128-3p | 1 |
| GO:0044788 | modulation by host of viral process | 0.039 | *PHB,NUCKS1* | viral processes | hsa-miR-26a-5p | 1 |
| GO:0072331 | signal transduction by p53 class mediator | 0.04 | *MDM2,CNOT4,CNOT2,RPL23* | cell cycle | hsa-miR-363-3p | 1 |
| GO:0097168 | mesenchymal stem cell proliferation | 0.041 | *CCNE1* | cell cycle | hsa-miR-144-5p | 1 |
| REAC:R-HSA-1630316 | Glycosaminoglycan metabolism | 0.041 | *HS3ST1* | hormone and metabolites | hsa-miR-144-5p | 1 |
| REAC:R-HSA-69206 | G1/S Transition | 0.041 | *CCNE1* | cell cycle | hsa-miR-144-5p | 1 |
| REAC:R-HSA-69306 | DNA Replication | 0.041 | *CCNE1* | cell cycle | hsa-miR-144-5p | 1 |
| REAC:R-HSA-196299 | Beta-catenin phosphorylation cascade | 0.041 | *GSK3B,PPP2R5D* | Wnt/beta-catenin signaling pathway | hsa-miR-26a-5p | 1 |
| REAC:R-HSA-6782135 | Dual incision in TC-NER | 0.041 | *RPS27A,DDB1,POLR2E* | cell cycle | hsa-miR-26a-5p | 1 |
| REAC:R-HSA-6782210 | Gap-filling DNA repair synthesis and ligation in TC-NER | 0.041 | *RPS27A,DDB1,POLR2E* | cell cycle | hsa-miR-26a-5p | 1 |
| REAC:R-HSA-6807070 | PTEN Regulation | 0.041 | *RPS27A,TNRC6B,RCOR1,MTA3* | cell cycle | hsa-miR-26a-5p | 1 |
| REAC:R-HSA-69563 | p53-Dependent G1 DNA Damage Response | 0.041 | *CCNE1,RPS27A,MDM2* | cell cycle | hsa-miR-144-5p, hsa-miR-144-5p, hsa-miR-26a-5p, hsa-miR-26a-5p | 4 |

| | | | | | | |
|---|---|---|---|---|---|---|
| REAC:R-HSA-69563 | p53-Dependent G1 DNA Damage Response | 0.041 | *CCNE1,RPS27A,MDM2* | cell cycle | hsa-miR-144-5p, hsa-miR-144-5p, hsa-miR-26a-5p, hsa-miR-26a-5p | 4 |
| REAC:R-HSA-69580 | p53-Dependent G1/S DNA damage checkpoint | 0.041 | *CCNE1,RPS27A,MDM2* | cell cycle | hsa-miR-144-5p, hsa-miR-144-5p, hsa-miR-26a-5p, hsa-miR-26a-5p | 4 |
| REAC:R-HSA-69580 | p53-Dependent G1/S DNA damage checkpoint | 0.041 | *CCNE1,RPS27A,MDM2* | cell cycle | hsa-miR-144-5p, hsa-miR-144-5p, hsa-miR-26a-5p, hsa-miR-26a-5p | 4 |
| GO:0032606 | type I interferon production | 0.041 | *IRAK1,IRF7* | cytokine | hsa-miR-146a-5p | 1 |
| REAC:R-HSA-5663202 | Diseases of signal transduction by growth factor receptors and second messengers | 0.041 | *GSK3B,RPS27A,PHB,MDM2,PPP2R5D,POLR2E* | diseases | hsa-miR-26a-5p | 1 |
| REAC:R-HSA-69615 | G1/S DNA Damage Checkpoints | 0.041 | *CCNE1,RPS27A,MDM2* | cell cycle | hsa-miR-144-5p, hsa-miR-144-5p, hsa-miR-26a-5p, hsa-miR-26a-5p | 4 |
| REAC:R-HSA-69615 | G1/S DNA Damage Checkpoints | 0.041 | *CCNE1,RPS27A,MDM2* | cell cycle | hsa-miR-144-5p, hsa-miR-144-5p, hsa-miR-26a-5p, hsa-miR-26a-5p | 4 |
| REAC:R-HSA-5250913 | Positive epigenetic regulation of rRNA expression | 0.041 | *GSK3B,MTA3,POLR2E* | epigenetic changes | hsa-miR-26a-5p | 1 |
| GO:0000377 | RNA splicing, via transesterification reactions with bulged adenosine as nucleophile | 0.041 | *HNRNPU,SFPQ,POLR2E,HNRNPA0,CPSF2* | transcription and splicing | hsa-miR-26a-5p | 1 |
| GO:0006289 | nucleotide-excision repair | 0.041 | *RPS27A,DDB1,POLR2E* | cell cycle | hsa-miR-26a-5p | 1 |
| REAC:R-HSA-6804760 | Regulation of TP53 Activity through Methylation | 0.041 | *RPS27A,MDM2* | epigenetic changes | hsa-miR-26a-5p | 1 |
| GO:0000375 | RNA splicing, via transesterification reactions | 0.042 | *HNRNPU,SFPQ,POLR2E,HNRNPA0,CPSF2* | transcription and splicing | hsa-miR-26a-5p | 1 |
| KEGG:04110 | Cell cycle | 0.043 | *GSK3B,CCNE1,YWHAE,MDM2* | cell cycle | hsa-miR-26a-5p | 1 |
| KEGG:04120 | Ubiquitin mediated proteolysis | 0.043 | *UBA2,RPS27A,DDB1,MDM2* | protein synthesis | hsa-miR-26a-5p | 1 |
| REAC:R-HSA-109606 | Intrinsic Pathway for Apoptosis | 0.044 | *PPP1R13B,AKT3* | apoptosis and senescence | hsa-miR-29c-3p | 1 |
| REAC:R-HSA-202131 | Metabolism of nitric oxide: NOS3 activation and regulation | 0.044 | *WASL,DDAH1* | cellular signaling | hsa-miR-128-3p | 1 |
| GO:0031109 | microtubule polymerization or depolymerization | 0.044 | *ZNF207,BLOC1S2,TUBGCP5* | cellular architecture | hsa-miR-26a-5p | 1 |
| GO:0044843 | cell cycle G1/S phase transition | 0.044 | *MDM2,CNOT4,CNOT2,FBXW7* | cell cycle | hsa-miR-363-3p | 1 |
| REAC:R-HSA-453279 | Mitotic G1 phase and G1/S transition | 0.044 | *CCNE1* | cell cycle | hsa-miR-144-5p | 1 |
| REAC:R-HSA-2995383 | Initiation of Nuclear Envelope (NE) Reformation | 0.045 | *LMNB1,KPNB1* | cell cycle | hsa-miR-128-3p | 1 |
| REAC:R-HSA-2559583 | Cellular Senescence | 0.045 | *CCNE1* | senescence | hsa-miR-144-5p, hsa-miR-26a-5p | 2 |
| REAC:R-HSA-69242 | S Phase | 0.045 | *CCNE1* | cell cycle | hsa-miR-144-5p | 1 |
| KEGG:04064 | NF-kappa B signaling pathway | 0.046 | *IRAK1,CARD10* | NF kappa beta signaling pathway | hsa-miR-146a-5p | 1 |

| | | | | | | |
|---|---|---|---|---|---|---|
| KEGG:04620 | Toll-like receptor signaling pathway | 0.046 | *IRAK1,IRF7* | toll like receptor signaling pathway | hsa-miR-146a-5p | 1 |
| KEGG:04933 | AGE-RAGE signaling pathway in diabetic complications | 0.046 | *SMAD2,PRKCE* | signaling activity | hsa-miR-146a-5p | 1 |
| KEGG:05142 | Chagas disease | 0.046 | *IRAK1,SMAD2* | diseases | hsa-miR-146a-5p | 1 |
| REAC:R-HSA-73854 | RNA Polymerase I Promoter Clearance | 0.047 | *UBTF,MTA3,POLR2E* | cell cycle | hsa-miR-26a-5p | 1 |
| REAC:R-HSA-73864 | RNA Polymerase I Transcription | 0.047 | *UBTF,MTA3,POLR2E* | cell cycle | hsa-miR-26a-5p | 1 |
| REAC:R-HSA-2559583 | Cellular Senescence | 0.047 | *CCNE1,RPS27A,TNRC6B,MDM2* | senescence | hsa-miR-144-5p, hsa-miR-26a-5p | 2 |
| REAC:R-HSA-1502540 | Signaling by Activin | 0.048 | *SMAD2* | hormones | hsa-miR-146a-5p | 1 |
| REAC:R-HSA-209543 | p75NTR recruits signalling complexes | 0.048 | *IRAK1* | cellular signaling | hsa-miR-146a-5p | 1 |
| REAC:R-HSA-209560 | NF-kB is activated and signals survival | 0.048 | *IRAK1* | nF kappa beta signaling pathway | hsa-miR-146a-5p | 1 |
| REAC:R-HSA-9013973 | TICAM1-dependent activation of IRF3/IRF7 | 0.048 | *IRF7* | cellular signaling | hsa-miR-146a-5p | 1 |
| REAC:R-HSA-918233 | TRAF3-dependent IRF activation pathway | 0.048 | *IRF7* | cellular signaling | hsa-miR-146a-5p | 1 |
| REAC:R-HSA-975144 | IRAK1 recruits IKK complex upon TLR7/8 or 9 stimulation | 0.048 | *IRAK1* | toll like receptor signaling pathway | hsa-miR-146a-5p | 1 |
| GO:0034504 | protein localization to nucleus | 0.048 | *GSK3B,HNRNPU,YWHAE,MDM2* | cellular architecture | hsa-miR-26a-5p | 1 |
| REAC:R-HSA-6781827 | Transcription-Coupled Nucleotide Excision Repair (TC-NER) | 0.048 | *RPS27A,DDB1,POLR2E* | transcription and splicing | hsa-miR-26a-5p | 1 |
| REAC:R-HSA-2995410 | Nuclear Envelope (NE) Reassembly | 0.05 | *LMNB1,KPNB1,TNPO1* | cell cycle | hsa-miR-128-3p | 1 |
| GO:0006296 | nucleotide-excision repair, DNA incision, 5'-to lesion | 0.05 | *RPS27A,DDB1* | cell cycle | hsa-miR-26a-5p | 1 |

**Supplementary table 6: Role of selected brain specific transcription factors for which binding is affected by significant SNPs associated with Aβ42, BACE1 and sTREM2 levels in the CSF**

| **Transcription factor** | **Clinical significance** |
|---|---|
| AP-1_known1 | upregulation of BDNF in cortical neurons of rats [1]; regulation of dendrite growth in the drosophilia [2]; involved in the regulation of APP in human glial cells [3] |
| CTCF_disc1, CTCF_known1 | increased APP expression in hippocampus of rat model [4]; in mice downregulation of CTCF transcription factor increased in the number of microglia in the anterior cingulate cortex leading to gliosis and eventually neuronal death [5] |
| Ets_known1 | regulates axon guidance and dendritic morphology during development [6] |
| Foxa_known4, Foxf1, Foxf2, Foxj2_2, Foxl1_1, Foxo_1, Foxo_2, Foxq1 | Forkhead transcription factors family are involved in neuronal death, neuronal response to amyloid ß exposure leading to mitochondrial dysfunction, pro-inflammatory cytokines production, apoptosis [7, 8] |
| GATA_known1 | regulates SCNA transcription in dopaminergic neurons leading to increased levels of α-synuclein [9] |
| Hbp1 | neuronal differentiation in neurogenesis in mice models [10] |
| HDAC2_disc3 | reduced expression of HDAC2-Sp3 in AD patients and AD mouse models, inhibition of HDAC2-Sp3 increased synaptic activity and plasticity in an AD mouse model [11] |
| Hoxa5_1 | motor neuron differentiation in mice models [12] |
| Hsf_disc1 | involved in proteostasis in neuronal cells through the regulation of HSP (heat shock protein) expression [13]; increases APP expression through binding of heat shock elements [14] |
| Mef2_known3 | involved in microglial homeostasis; suppression of Mef2 associated with microglial phenotype associated with amyloid plaques in AD cortex [15] |
| Mxi1_disc1 | involved in neuronal differentiation in xenopus [16] |
| NRSF_known1, NRSF_known2, NRSF_known3 | NRSF (also known as repression element 1 silencing transcription factor) bind to neuron-restrictive silencer elements of the choline acetyltransferase gene in non-neuronal cells of animal models [17, 18]; protective effect on neuronal cells [19]; missense variation in REST associated with hippocampal volume loss [20]; |
| PLZF | involved in cortical neurogenesis in mouse brains [21]; interacts with CEBPD to inhibit apoptosis in astrocytes of AD mice models [22] |
| Pou5f1_disc1, Pou5f1_known2 | involved in cortical neurogenesis in mice [23] |
| RXRA_known4 | variation in RXRa gene increases risk of AD through the regulation of genes involved in cholesterol metabolism [24] |
| Sin3Ak-20_disc7 | regulated the expression of neuronal genes and neuronal differentiation [25] |
| Sox_2 | stimulates the non-amyloidogenic processing of ßAPP by stimulating ADAM10 activity [26] |
| SP1_disc3 | regulates expression of APP, tau, PSEN2 promoter transcription and BACE1 [27-32]; in AD mouse model, inhibition of Sp1 increased memory deficits [33] |

| | |
|---|---|
| SP2_disc3 | involved in neurogenesis [34] |
| SREBP_disc1 | regulates cholesterol metabolism for the synthesis of the myelin membrane [35] |
| TAL1_disc1 | involved in neurogenesis of GABAergic neurons [36] |
| TCF12_disc2, | involved in neurogenesis in mice models [37] |
| YY1_disc2, YY1_known6 | involved in neurogenesis in mice models [38] |

**References**


[1] Tuvikene J, Pruunsild P, Orav E, Esvald EE, Timmusk T. AP-1 Transcription Factors Mediate BDNF-Positive Feedback Loop in Cortical Neurons. The Journal of neuroscience : the official journal of the Society for Neuroscience. 2016;36:1290-305.
[2] Hartwig CL, Worrell J, Levine RB, Ramaswami M, Sanyal S. Normal dendrite growth in Drosophila motor neurons requires the AP-1 transcription factor. Developmental neurobiology. 2008;68:1225-42.
[3] Trejo J, Massamiri T, Deng T, Dewji NN, Bayney RM, Brown JH. A direct role for protein kinase C and the transcription factor Jun/AP-1 in the regulation of the Alzheimer's beta-amyloid precursor protein gene. The Journal of biological chemistry. 1994;269:21682-90.
[4] Yang Y, Quitschke WW, Vostrov AA, Brewer GJ. CTCF is essential for up-regulating expression from the amyloid precursor protein promoter during differentiation of primary hippocampal neurons. Journal of neurochemistry. 1999;73:2286-98.
[5] Kwak J-H, Kim S, Yu N-K, Seo H, Choi JE, Kim J-i, et al. Loss of the neuronal genome organizer and transcription factor CTCF induces neuronal death and reactive gliosis in the anterior cingulate cortex. Genes, Brain and Behavior. 2020;n/a:e12701.
[6] Santiago C, Bashaw GJ. Transcription factors and effectors that regulate neuronal morphology. Development (Cambridge, England). 2014;141:4667-80.
[7] Maiese K. Forkhead Transcription Factors: Formulating a FOXO Target for Cognitive Loss. Current neurovascular research. 2017;14:415-20.
[8] Maiese K. Forkhead transcription factors: new considerations for alzheimer's disease and dementia. Journal of translational science. 2016;2:241-7.
[9] Scherzer CR, Grass JA, Liao Z, Pepivani I, Zheng B, Eklund AC, et al. GATA transcription factors directly regulate the Parkinson's disease-linked gene α-synuclein. Proceedings of the National Academy of Sciences. 2008;105:10907.
[10] Watanabe N, Kageyama R, Ohtsuka T. Hbp1 regulates the timing of neuronal differentiation during cortical development by controlling cell cycle progression. Development (Cambridge, England). 2015;142:2278-90.
[11] Yamakawa H, Cheng J, Penney J, Gao F, Rueda R, Wang J, et al. The Transcription Factor Sp3 Cooperates with HDAC2 to Regulate Synaptic Function and Plasticity in Neurons. Cell Reports. 2017;20:1319-34.

[12] Catela C, Shin MM, Lee DH, Liu JP, Dasen JS. Hox Proteins Coordinate Motor Neuron Differentiation and Connectivity Programs through Ret/Gfrα Genes. Cell reports. 2016;14:1901-15.
[13] Qu Z, Titus ASCLS, Xuan Z, D'Mello SR. Neuroprotection by Heat Shock Factor-1 (HSF1) and Trimerization-Deficient Mutant Identifies Novel Alterations in Gene Expression. Scientific Reports. 2018;8:17255.
[14] Theuns J, Van Broeckhoven C. Transcriptional regulation of Alzheimer's disease genes: implications for susceptibility. Human Molecular Genetics. 2000;9:2383-94.
[15] Krasemann S, Madore C, Cialic R, Baufeld C, Calcagno N, El Fatimy R, et al. The TREM2-APOE Pathway Drives the Transcriptional Phenotype of Dysfunctional Microglia in Neurodegenerative Diseases. Immunity. 2017;47:566-81.e9.
[16] Klisch TJ, Souopgui J, Juergens K, Rust B, Pieler T, Henningfeld KA. Mxi1 is essential for neurogenesis in Xenopus and acts by bridging the pan-neural and proneural genes. Developmental biology. 2006;292:470-85.
[17] Shimojo M, Paquette AJ, Anderson DJ, Hersh LB. Protein kinase A regulates cholinergic gene expression in PC12 cells: REST4 silences the silencing activity of neuron-restrictive silencer factor/REST. Mol Cell Biol. 1999;19:6788-95.
[18] Hersh LB, Shimojo M. Regulation of cholinergic gene expression by the neuron restrictive silencer factor/repressor element-1 silencing transcription factor. Life Sci. 2003;72:2021-8.
[19] Mozzi A, Guerini FR, Forni D, Costa AS, Nemni R, Baglio F, et al. REST, a master regulator of neurogenesis, evolved under strong positive selection in humans and in non human primates. Scientific Reports. 2017;7:9530.
[20] Nho K, Kim S, Risacher SL, Shen L, Corneveaux JJ, Swaminathan S, et al. Protective variant for hippocampal atrophy identified by whole exome sequencing. Annals of neurology. 2015;77:547-52.
[21] Lin H-C, Ching Y-H, Huang C-C, Pao P-C, Lee Y-H, Chang W-C, et al. Promyelocytic leukemia zinc finger is involved in the formation of deep layer cortical neurons. Journal of Biomedical Science. 2019;26:30.
[22] Wang SM, Lee YC, Ko CY, Lai MD, Lin DY, Pao PC, et al. Increase of zinc finger protein 179 in response to CCAAT/enhancer binding protein delta conferring an antiapoptotic effect in astrocytes of Alzheimer's disease. Mol Neurobiol. 2015;51:370-82.
[23] McEvilly RJ, de Diaz MO, Schonemann MD, Hooshmand F, Rosenfeld MG. Transcriptional regulation of cortical neuron migration by POU domain factors. Science (New York, NY). 2002;295:1528-32.
[24] Kölsch H, Lütjohann D, Jessen F, Popp J, Hentschel F, Kelemen P, et al. RXRA gene variations influence Alzheimer's disease risk and cholesterol metabolism. Journal of cellular and molecular medicine. 2009;13:589-98.
[25] Chaubal A, Pile LA. Same agent, different messages: insight into transcriptional regulation by SIN3 isoforms. Epigenetics & Chromatin. 2018;11:17.
[26] Sarlak G, Htoo HH, Hernandez JF, Iizasa H, Checler F, Konietzko U, et al. Sox2 functionally interacts with βAPP, the βAPP intracellular domain and ADAM10 at a transcriptional level in human cells. Neuroscience. 2016;312:153-64.
[27] Lukiw WJ, Rogaev EI, Wong L, Vaula G, McLachlan DR, St George Hyslop P. Protein-DNA interactions in the promoter region of the amyloid precursor protein (APP) gene in human neocortex. Brain research Molecular brain research. 1994;22:121-31.

[28] Querfurth HW, Jiang J, Xia W, Selkoe DJ. Enhancer function and novel DNA binding protein activity in the near upstream betaAPP gene promoter. Gene. 1999;232:125-41.
[29] Docagne F, Gabriel C, Lebeurrier N, Lesné S, Hommet Y, Plawinski L, et al. Sp1 and Smad transcription factors co-operate to mediate TGF-beta-dependent activation of amyloid-beta precursor protein gene transcription. The Biochemical journal. 2004;383:393-9.
[30] Christensen MA, Zhou W, Qing H, Lehman A, Philipsen S, Song W. Transcriptional regulation of BACE1, the beta-amyloid precursor protein beta-secretase, by Sp1. Molecular and cellular biology. 2004;24:865-74.
[31] Heicklen-Klein A, Ginzburg I. Tau promoter confers neuronal specificity and binds Sp1 and AP-2. Journal of neurochemistry. 2000;75:1408-18.
[32] Renbaum P, Beeri R, Gabai E, Amiel M, Gal M, Ehrengruber MU, et al. Egr-1 upregulates the Alzheimer's disease presenilin-2 gene in neuronal cells. Gene. 2003;318:113-24.
[33] Citron BA, Saykally JN, Cao C, Dennis JS, Runfeldt M, Arendash GW. Transcription factor Sp1 inhibition, memory, and cytokines in a mouse model of Alzheimer's disease. American journal of neurodegenerative disease. 2015;4:40-8.
[34] Liang H, Xiao G, Yin H, Hippenmeyer S, Horowitz JM, Ghashghaei HT. Neural development is dependent on the function of specificity protein 2 in cell cycle progression. Development (Cambridge, England). 2013;140:552-61.
[35] Camargo N, Smit AB, Verheijen MHG. SREBPs: SREBP function in glia–neuron interactions. The FEBS Journal. 2009;276:628-36.
[36] Achim K, Peltopuro P, Lahti L, Tsai HH, Zachariah A, Astrand M, et al. The role of Tal2 and Tal1 in the differentiation of midbrain GABAergic neuron precursors. Biology open. 2013;2:990-7.
[37] Uittenbogaard M, Chiaramello A. Expression of the bHLH transcription factor Tcf12 (ME1) gene is linked to the expansion of precursor cell populations during neurogenesis. Brain research Gene expression patterns. 2002;1:115-21.
[38] Zurkirchen L, Varum S, Giger S, Klug A, Häusel J, Bossart R, et al. Yin Yang 1 sustains biosynthetic demands during brain development in a stage-specific manner. Nature Communications. 2019;10:2192.
